# Factors Associated with Divergent Introgression Outcomes from *Wolbachia*-Based Interventions in Rio de Janeiro, Brazil

**DOI:** 10.64898/2026.10.16.26365711

**Authors:** Elisabeth Nelson, Paloma M. Cárcamo, Gabriel Ribeiro dos Santos, Gabriel Sylvestre Ribeiro, Thiago Rodrigues da Costa, Ilaria Dorigatti, Henrik Salje, Luciano A. Moreira, Derek A.T. Cummings, Albert I. Ko

**Author notes:** Authors contributed equally (Elisabeth Nelson, Paloma M. Cárcamo, Luciano A. Moreira, Derek A.T. Cummings, Albert I. Ko).

## Abstract

*Wolbachia*-based interventions were less effective in achieving introgression in Rio de Janeiro than other settings. Reduced introgression occurred in locations where dengue was historically high, raising concerns of inequity. We examined factors affecting *Wolbachia* introgression in Rio through trajectory modeling, spatiotemporal Bayesian models, and *Ae. aegypti* abundance estimation. During the intervention (2017-2020), *Wolbachia* introgression was 26% (median; IQR,14-44%), with four distinct trajectories. Despite receiving more releases, densely populated areas with low socioeconomic status had lower introgression. In spatiotemporally explicit multivariable regressions, higher *Ae. aegypti* abundance, temperature, population density and lower socioeconomic status were associated with reduced introgression. Improved characterization of *Ae. aegypti* abundance and other structural/environmental predictors of introgression could be used to calibrate release doses to ensure equitable benefits from *Wolbachia* interventions.

## Main Text

The global burden of dengue and other arboviral diseases has been increasing worldwide, highlighting the need for effective interventions to control transmission (1,2). Transinfection of the endosymbiotic bacteria *Wolbachia pipientis* in *Aedes aegypti* mosquitoes, the main vector for dengue, can block transmission of dengue virus (DENV) and other arboviruses within the mosquito (3,4). *Wolbachia* infection induces cytoplasmic incompatibility in *Ae. aegypti*, meaning that uninfected females that mate with infected males produce infertile eggs, while infected females will have viable eggs. This phenomenon allows for *Wolbachia* infection to rapidly and sustainably introgress and remain stable in *Ae. aegypti* populations over long periods if sufficient numbers of *Wolbachia* infected mosquitoes are released into an area resulting in increased frequency-dependent fitness of infected females compared to uninfected mosquitoes (5,6).

The potential of *Wolbachia*-based interventions to reduce arboviral disease transmission has led to several studies and interventions. A randomized trial conducted in Yogyakarta found that after achieving high levels of introgression, i.e. prevalence, of the *w*Mel strain of *Wolbachia* (∼98.5%), dengue risk was reduced by approximately 77% (7). However, in other settings, such as Rio de Janeiro in Brazil, releases have been less effective at achieving high levels of introgression and reducing risk of arboviral disease (8). In addition, the effectiveness of the intervention in this setting was found to be highly heterogeneous across release areas. Areas with historically higher rates of dengue and chikungunya incidence achieved lower levels of introgression, and therefore, lower reductions in arboviral disease. Our study aimed to characterize the variability in introgression in Rio de Janeiro and quantify the impact of operational, meteorological, socioeconomic and environmental factors on introgression to inform optimization of *w*Mel releases and achieve equitable introgression.

### Implementation of *w*Mel releases and introgression monitoring in Rio de Janeiro

The World Mosquito Program (WMP) released adult *w*Mel-positive *Ae. aegypti* in an 86.8 km^2^ area in northwest Rio de Janeiro (**Figure 1A**) between August 2017 and March 2020 (8,9). The area was divided into five intervention sites, which followed different release schedules. Tubes containing an average of 350 mosquitoes were emptied every 50m by vehicle or by foot. *w*Mel introgression was monitored using a network of 1168 BG-traps, which were collected biweekly. Up to 10 *Ae. aegypti* per trap were tested for *w*Mel using a validated qPCR assay. Introgression was estimated as the proportion of tested mosquitoes that were positive for *w*Mel (i.e. *w*Mel prevalence among host-seeking adult females captured in BG-Sentinel traps). If a trap was found to have consistently low levels of introgression, releases were continued in the surrounding area, leading to differing lengths of the intervention across the study area (**Figure 1D**).

**Figure 1.**
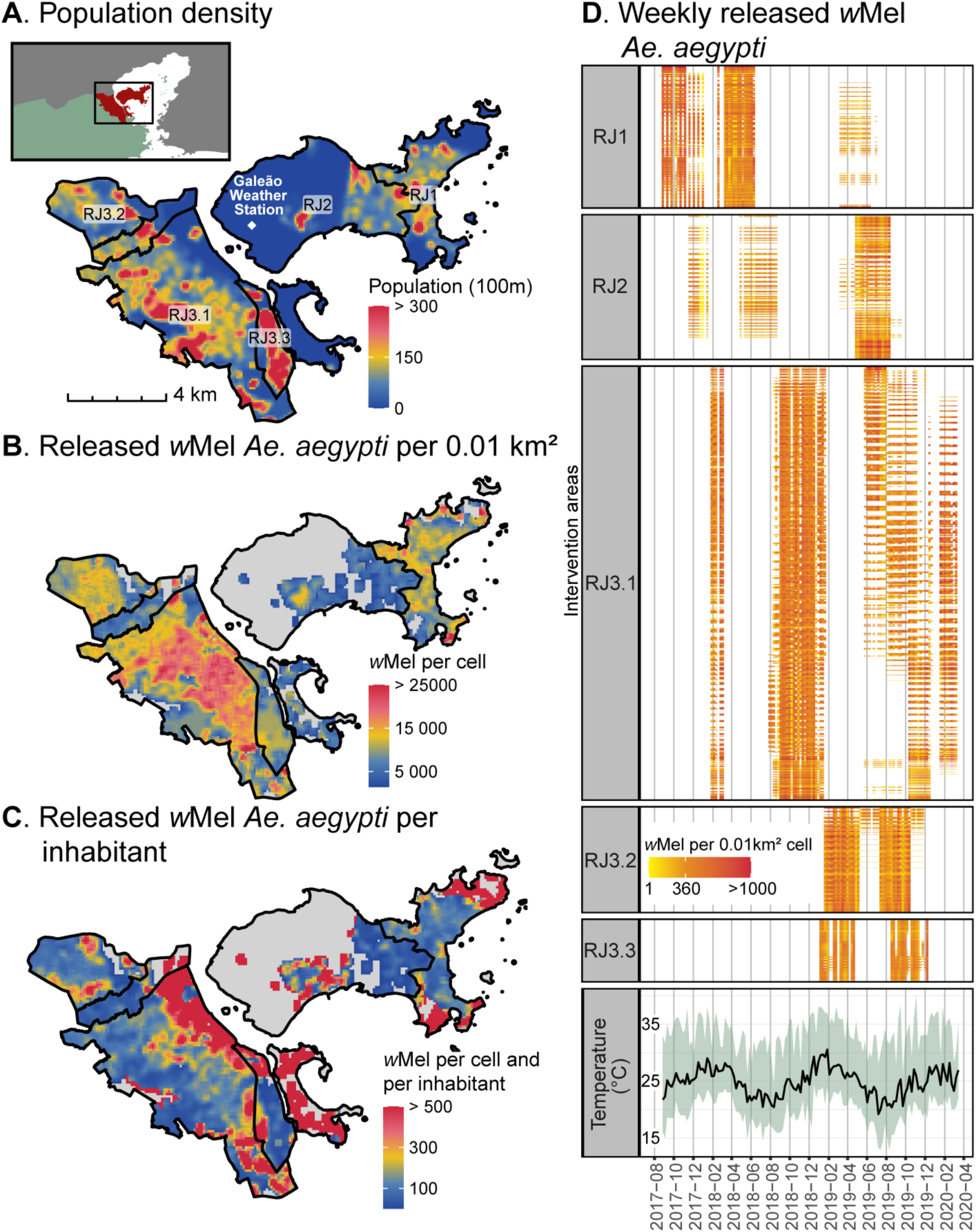
Roll-out of *w*Mel intervention in Rio de Janeiro, 2017-2020. (A, B, C) Maps of population density and releases. The intervention was not implemented in areas shaded in gray. (D) Release schedule of each intervention area. Rows represent 0.01km^2^ cells. Daily temperature (mean, minimum and maximum) in degrees Celsius measured in Galeão Station, located in RJ2.

By the end of the intervention, there was marked heterogeneity in the cumulative dose of released *w*Mel *Ae. aegypti* across the intervention area (median 10,428, interquartile range [IQR] 6,854-14,740 mosquitoes per 0.01km^2^) (**Figure 1B**), as well as in the cumulative dose of released *w*Mel *Ae. aegypti* per inhabitant (median 79.10, IQR 21.26-171.57 mosquitoes per inhabitant in 0.01km^2^) (**Figure 1C**). Areas with lower cumulative dose per inhabitant had lower income levels and less impervious surfaces (**Tables S1, S2**).

### Characterization of trap-level introgression trajectories through time

Mean *w*Mel introgression throughout the study period was low in most traps (median 0.26, IQR 0.14-0.44) (**Figure 2A**). We conducted group-based trajectory modeling on spline-smoothed trap-level introgression trajectories and identified four clusters of distinct patterns of introgression trajectories (**Figures 2B-C**). We observed an inverse association between introgression and cumulative releases at the cluster level, suggesting that continued releases of the same dose over time may not be enough to ensure introgression. The cluster with the most successful introgression (Cluster 1) had received the lowest cumulative dose of *w*Mel *Ae. aegypti* releases by area, while the cluster with lowest mean introgression (Cluster 4) had received the highest cumulative releases (**Figure 2D**).

**Figure 2.**
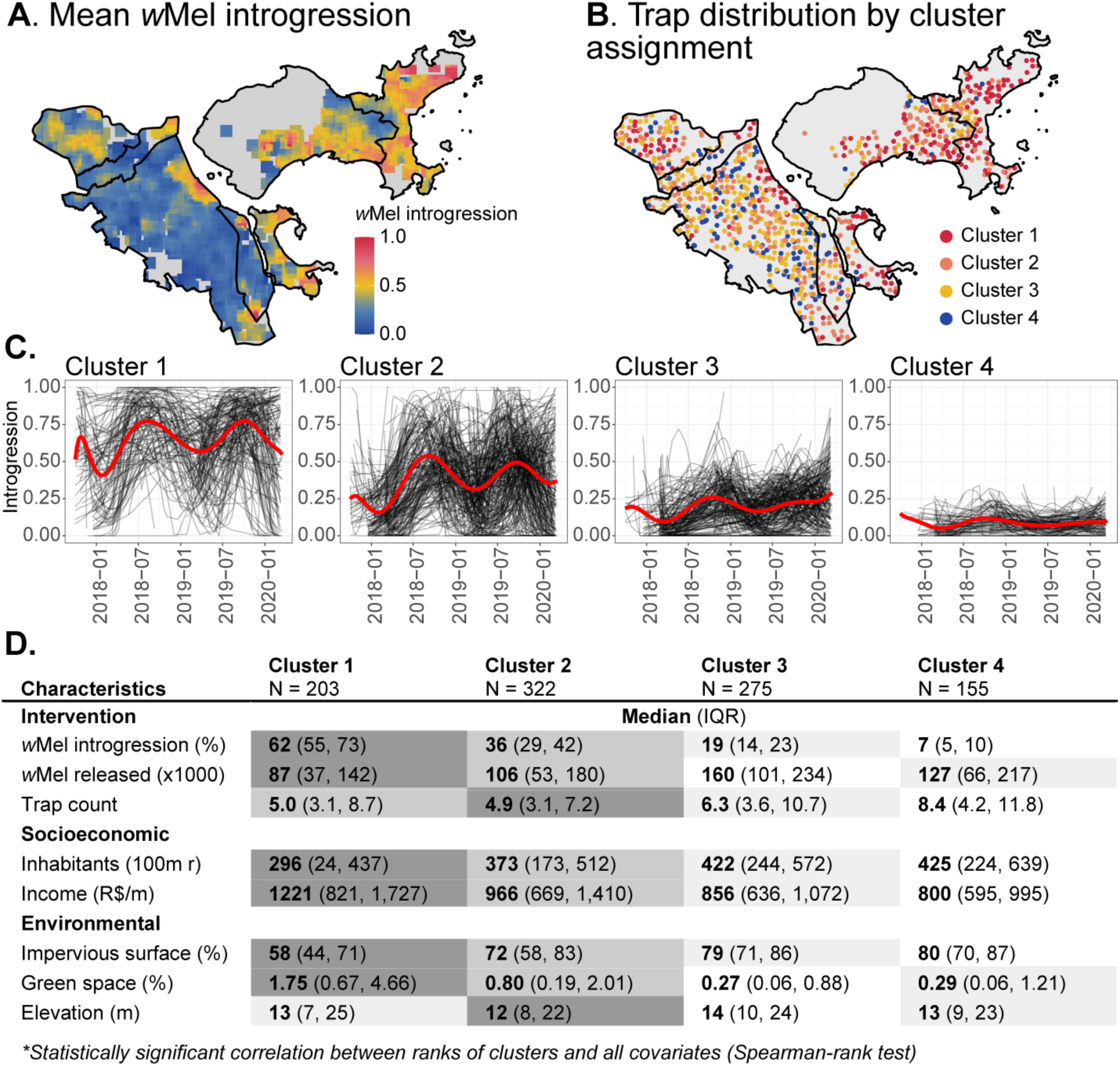
Group-based multivariate trajectory modeling (GBMT) of *w*Mel introgression trajectories. (A) Map of mean *w*Mel introgression throughout the study period. Gray areas did not have traps. (B) Map of spatial distribution of 955 traps included in GBMT. (C) Smoothed introgression trajectories of individual traps stratified by cluster. Red lines are the estimated trajectory for each cluster. (D) Characteristics of intervention, socioeconomic, and environmental factors in 100m radius of each trap. Boxes are shaded according to each cluster’s rank for a specific characteristic.

To explore factors that could explain the observed differences in introgression trajectories, we obtained total *Ae. aegypti* counts from each trap and extracted information on socioeconomic and environmental covariates from a 100m radius around each trap. We found that clusters with highest introgression had lower mosquito abundance as measured by *Ae. aegypti* trap counts (**Figure 2D**). We also observed that clusters with higher levels of introgression had lower human population density, higher income, less impervious surfaces and more green spaces (**Table S3**).

### Quantification of spatiotemporal factors influencing trap-level introgression

To formally test the influence of operational, meteorological, socioeconomic, environmental, and entomological variables on introgression, we developed a spatiotemporally explicit multivariable model using the Integrated Nested Laplace Approximation (INLA) method of Bayesian inference. Using trap-level *w*Mel prevalence as the outcome, we performed a hierarchical model fitting process, successively testing a wide range of covariates for association with the outcome (**Table S4**). Covariates significantly associated with the outcome which improved the model fit were included as either linear or non-linear depending on their observed association with the outcome, using univariate generalized additive models with a cubic regression spline to infer linearity (**Figure S1**). We expect environmental factors to impact different stages of the mosquito life cycle and may have delayed effects on abundance and introgression. To account for this, we assessed different temporal lags and selected the ones significantly correlated with the outcome in univariate models to use in the model (**Figure S2 and S3**).

We found that measures of *Ae. aegypti* abundance, previous introgression, meteorological conditions, socioeconomic indicators, and metrics of the built environment, were associated with lower introgression (**Figure 3**). Several of these factors are associated with larger *Ae. aegypti* populations. To account for potential contamination of introgression estimates during and directly after releases, we included terms related to whether releases occurred and the number of mosquitoes released in the previous week, which controls for the potential recapture of released *w*Mel-infected mosquitoes. We found increasing *Ae. aegypti* abundance had a negative association with introgression. We found the highest quintile of mosquito trap counts (27 *Ae. aegypti*) one-and two-weeks prior were associated with a 5% (95% CrI 2-8%) decrease in introgression as compared to the median trap count (4 *Ae. aegypti*). Increasing previous introgression one-and two-weeks prior were strongly associated with an increase in introgression by 76% (95% CrI 68-84%) and 34% (95% CrI 29-40%) respectively. It is likely that this autocorrelation captures the frequency-dependent fitness effects associated with *w*Mel introgression (cytoplasmic incompatibility and maternal inheritance). Notably, in a model excluding the lagged-introgression and abundance terms (**Figure S4**), the socioeconomic, environmental, and meteorological associations persisted with the same direction of effect. This indicates that these effects are not artifacts of the autoregressive structure; they are identified primarily from spatial (between-location) variation, which the temporal autoregressive term does not absorb.

**Figure 3.**
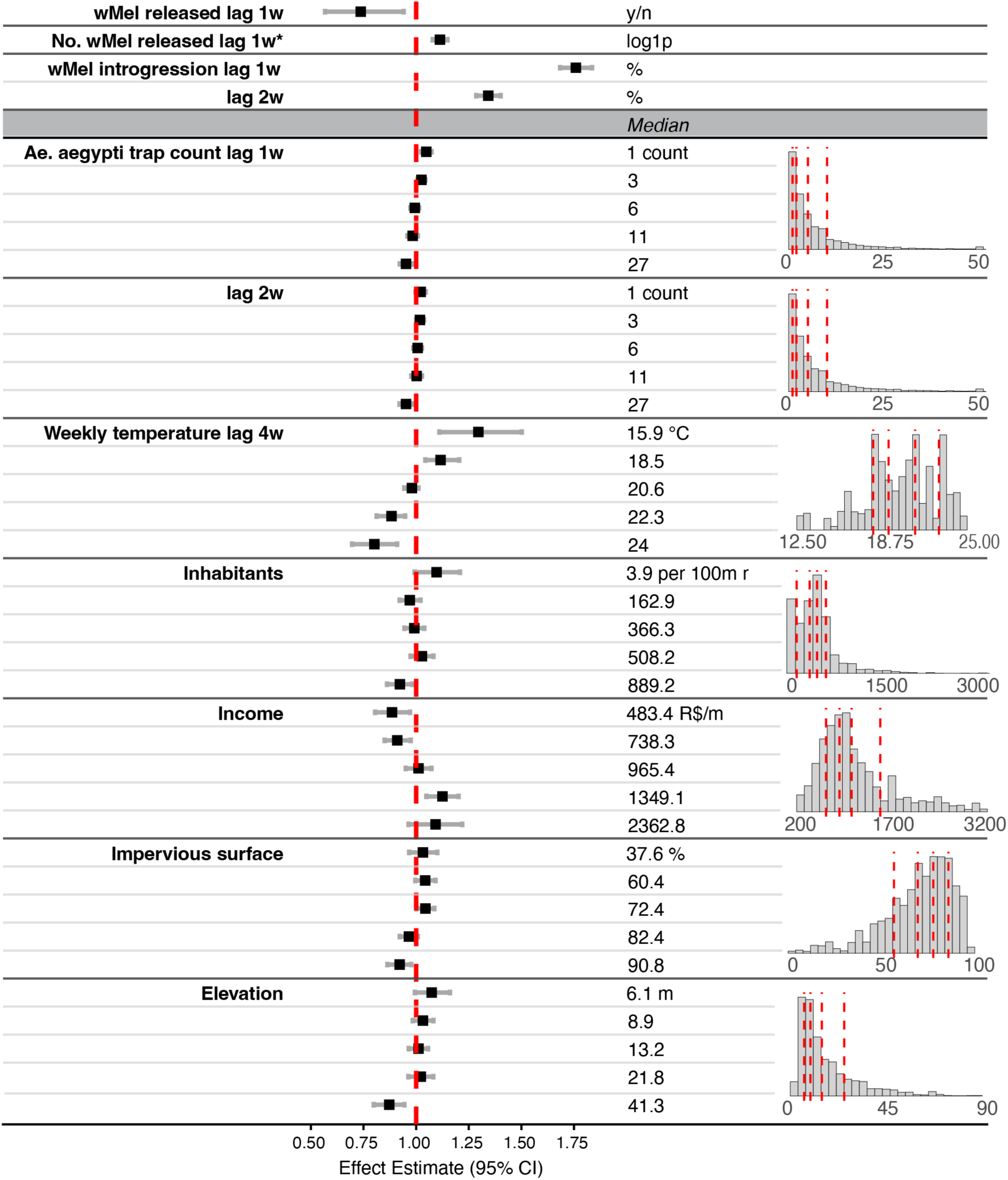
Spatiotemporal analysis of factors measured during the intervention associated with *w*Mel introgression. (Conditional R^2^=0.76). The outcome is extrapolated *w*Mel count (*w*Mel prevalence\**Ae. aegypti* caught). Terms above the gray ribbon were included as linear and those below as non-linear. Non-linear effects were run as quintiles, depicted in the distribution plots, with red lines marking the boundaries, and medians and units of measurement in the text column. Data was aggregated by week and scaled to the 100m radius surrounding traps. ‘*w*Mel released lag 1w’ is a binary term, indicating whether releases occurred; ‘No. *w*Mel released lag 1w*’ is the log(1+x) or log1p number of mosquitoes released; ‘*w*Mel introgression lag 1w, 2w’ is the percent *w*Mel-positive mosquitoes; ‘Daily precipitation lag 6w’ is the mean daily precipitation, in centimeters; ‘*Ae. aegypti* trap count lag 1w, 2w’ are measures of *Ae. aegypti* abundance, distribution tails not depicted; ‘Weekly temperature lag 4w’ is the minimum weekly temperature, in Celsius; ‘Inhabitants’ is the population density; ‘Income’ is the mean income in R$ for persons over 10; ‘Impervious surface’ is the percent area comprised of water-impervious surfaces; ‘Elevation’ is in meters.

Investigating the effects of temporally-varying meteorological factors, we found increasing minimum weekly temperature four weeks prior were negatively associated with introgression. Observations in the lowest quintile of minimum temperatures, around 15.9℃, had 29% (95% CrI 10-51%) more introgression than those with the median minimum temperature (20℃), while the highest quintile of minimum temperature, around 24℃, had a 20% (95% CrI 8-31%) negative effect relative to the median. Increasing minimum temperature four weeks prior is beneficial for the development of the previous generation of *Ae. aegypti* and for the egg and larval stages of the current adult population, having an indirect association with larger current *Ae. aegypti* populations (10–13). These temperature effects appear to be driven by seasonal temperature changes, rather than weather extremes (**Figure S5**). This suggests that introgression is less effective under conditions associated with larger wildtype *Ae. aegypti* populations, such as warmer seasons (5,14–16). Higher temperatures may also lead to lower introgression due to the degradation of *w*Mel *Wolbachia* in *Ae. aegypti* eggs at higher temperatures (17–20). The covariate selection process is fully documented in Table S4, and the consistency of associations across multiple sensitivity analyses (varying spatial scale, temporal autocorrelation structure, inclusion/exclusion of abundance terms, and restriction to non-release periods) provides empirical reassurance that the reported associations are not artifacts of the selection procedure.

Additionally, spatially varying factors associated with greater percentages of built environment and lower socioeconomic status (SES) were also negatively associated with introgression. Observations in the highest population density of around 889.2 inhabitants per 100m radius had 8% (95% CrI 1-14%) less introgression than the median population density (366.3 inhabitants per 100m radius). Population density is also often an indicator of *Ae. aegypti* abundance, with higher population density found to be associated with increased *Ae. aegypti* populations in other settings (21). Of note, *Ae. albopictus* is present in Rio de Janeiro but is largely restricted to peri-urban and forested areas rather than the dense urban neighborhoods covered by our study, making interspecific competition an unlikely major driver of the observed heterogeneity in introgression. Similarly, lowest income quintile (R$483.4 per month), had 12% (95% CrI 3-20%) less introgression than those with median income (R$964.5), whereas those in the second highest quintile (R$1349.1 per month), had 12% (95% CrI 5-20%) higher introgression than the median. Lower mean monthly income and other indicators of lower SES have been found to be associated with increased numbers of mosquito breeding sites and increased mosquito-human interactions (22). Observations at the highest elevation, around 41.3m, had 13% (95% CrI 5-20%) less introgression than those with median elevation (13.2m). In Rio de Janeiro, urban informal settlements are located at higher elevations, thus SES declines with increasing elevation. Observations in the highest impervious surface quintile, of 90.8%, had 8% (95% CrI 1-14%) less introgression than those with the median impervious surface (72.4%).

Introgression was better achieved in places with intermediate levels of impervious surfaces. Places with low levels of impervious surface could be less hospitable habitats for *Ae. aegypti* populations, while places with high levels of impervious surfaces could facilitate the proliferation of cryptic breeding sites and the presence of larger *Ae. aegypti* populations (13). Details on the fit of spatial and temporal autoregressive terms for both models and results from sensitivity analyses can be found in the supplements (**Figures S6-S13**), which yielded trends consistent with our main models.

Despite the trends in factors negatively associated with introgression being associated with larger *Ae. aegypti* populations, including *Ae. aegypti* abundance in the model does not entirely account for their effects. We separately assessed models that did not include mosquito monitoring data and used only information that could be collected prior to a *w*Mel release intervention and readily measured in many locales, i.e. meteorological conditions, socioeconomic indicators and metrics of the built environment (**Figure S4**). Comparison of the model results showed inclusion of *Ae. aegypti* abundance and previous *w*Mel introgression yielded a slight dilution in the effects of the meteorological, environmental, and socioeconomic factors. However, these factors still showed significant effects on introgression, suggesting that *Ae. aegypti* population size partially mediates the influence of these effects, but does not entirely account for them. Therefore, these factors may be acting independently through mechanisms outside of wildtype population size, in addition to their effects on population size. We note that our models use the extrapolated *w*Mel-positive count with an offset for the total *Ae. aegypti* captured, rather than the raw prevalence, so the inverse abundance association is unlikely to arise mechanically from a shared prevalence denominator; we therefore interpret it as reflecting reduced establishment rather than denominator inflation, while acknowledging that a residual mechanical contribution cannot be fully excluded. Additionally, it may also be due to trap captures imprecisely measuring *Ae. aegypti* populations, either due to sparse spatial sampling or variation in attractiveness of traps in different locations. We therefore investigated the potential use of more accurate measures of abundance.

### Prediction of introgression utilizing baseline entomological observations

We explored predictors of introgression that could be measured at the onset of the releases. To obtain a more precise estimate of *Ae. aegypti* abundance in the release areas, we used mark release recapture methods traditionally used for abundance measurements in ecology (23). We calculated the Fisher-Ford (FF) Index assuming the ratio of *w*Mel-infected to wild-type *Ae. aegypti* captured in traps reflected the ratio of *w*Mel mosquitoes released to the ambient wild-type population (23). We could only calculate this index for the first trap collection and if the collection occurred 36 days or less after the first release, when the majority of captured *w*Mel-positive mosquitoes are expected to have been released and not introgressed. We found a negative association between the baseline *Ae. aegypti* abundance and mean *w*Mel introgression. Conversely, the ratio of *w*Mel mosquitoes released to abundance had a positive association with mean *w*Mel introgression (**Figure 4**). Both associations were consistent across assumed daily survival probabilities from 0.75 to 0.90 (**Figure S14**). This release-to-abundance ratio was associated with whether mean introgression throughout the intervention was above 20% (**Figure S15-S16**). In addition, we found that higher percentages of green space (odds ratio [OR] 1.47 per standard deviation [SD] increase) and higher mean income of head of household (OR 1.52 per SD) were positively associated with mean *w*Mel introgression. In contrast, a higher percentage of impervious surfaces was negatively associated with mean introgression (OR 0.63 per SD).

**Figure 4.**
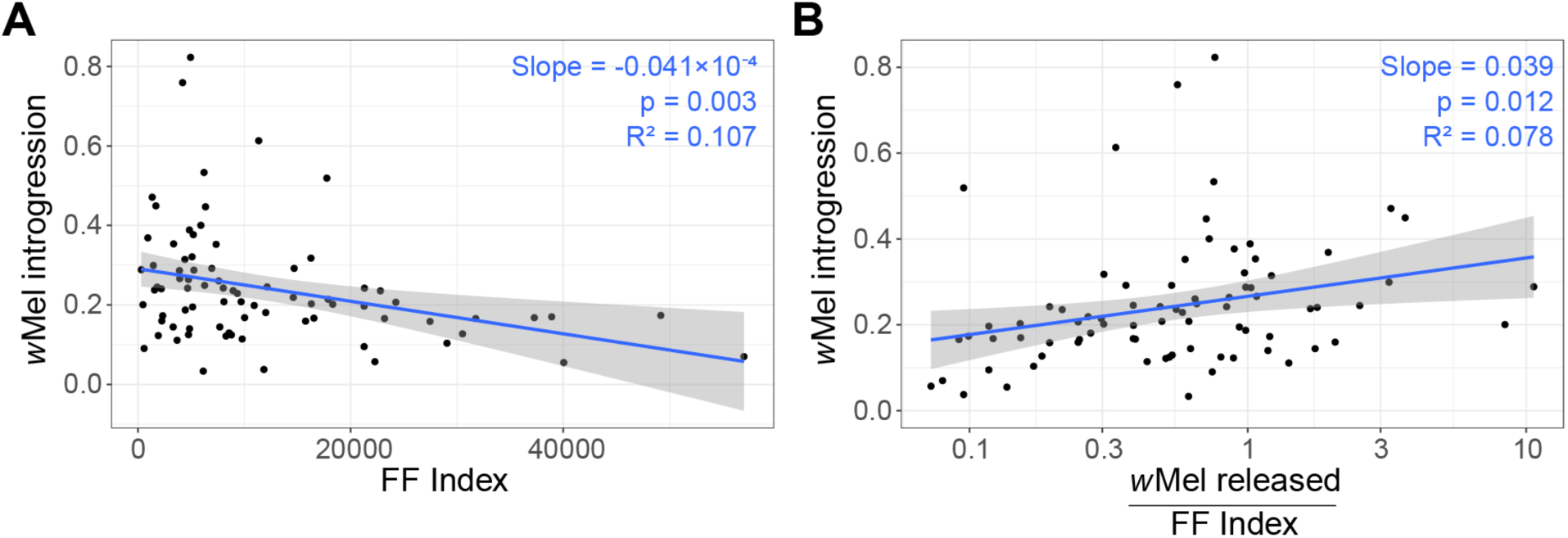
**Baseline abundance estimates and introgression in traps with 9-36 days between first release and first trapping event**. (A) Fisher-Ford (FF) Index and (B) ratio of wMel released (mean of releases before first trapping event) to Fisher-Ford Index versus mean *w*Mel introgression (log-transformed) for 85 traps; blue lines show linear regression trend with 95% CI. Blue text indicates regression slope, significance, and explained deviance (R^2^).

These findings suggest that baseline *Ae. aegypti* abundance, the relative dose of *w*Mel *Ae. aegypti* released compared to existing mosquito populations, landscape features and socioeconomic characteristics determine the success of the intervention. *Wolbachia* introgression has been described to achieve population replacement only if the proportion of *Wolbachia*-positive mosquitoes exceeds a certain threshold; introgression declines to zero otherwise (6). In our study setting, areas with high mosquito abundance may have not received large enough *w*Mel *Ae. aegypti* doses per release to exceeding this threshold and achieving population replacement, even if releases continued over a longer time. More robust estimates of baseline *Ae. aegypti* abundance will be helpful in confirming these associations and to better inform planning of releases. These baseline estimates rest on a limited number of traps (n = 85) and assume a fixed daily survival probability; immigration of wild-type mosquitoes from untreated areas and higher catchability of recently released compared to wild mosquitoes would both lead to underestimation of the FF index and therefore overestimation of the release-to-abundance ratio. We therefore present this analysis as hypothesis-generating and requiring confirmation through additional analyses.

## Limitations

There are several limitations inherent to our study. The number of mosquitoes caught during each trapping event was highly variable and introgression estimates were often based on the results of testing fewer than ten mosquitoes, which could lead to imprecise estimates. To address this, we smoothed introgression estimates, weighing by the number of mosquitoes tested, to account for differing levels of precision in different trapping events and used extrapolated *w*Mel counts as the outcome in our models.

Furthermore, our selection of covariates was constrained by the availability of data. We did not include spatially varying temperature and precipitation because there is only one Brazilian National Institute of Meteorology weather station in the study area. Nevertheless, given the small size of the study area, we do not expect significant spatial variability in these meteorological factors. Additionally, this weather station does not measure humidity, which has been shown to impact *Ae. aegypti* survival (24,25). We used BG-trap captures as a proxy for *Ae. aegypti* abundance, which may be underestimated in high-density, low-SES locations due to competition with human hosts for trap attractants. We did not include information on vector control practices or insecticide use due to lack of data availability. A change in the larvicide used in routine larval source management has been found in a recent paper to be associated with declines in long-term introgression stability in part of the intervention area, potentially due to *Wolbachia*-related fitness costs. (26). However, in our study, we found introgression levels remained stable through time at both high and low levels. Nonetheless, we included most factors shown in the literature to have a potential effect on *Ae. aegypti* abundance and/or *w*Mel introgression. Our socioeconomic indicators were derived from the 2010 census and thus predate the intervention by several years, and elevation served in part as a proxy for socioeconomic status; since these indicators are correlated with *Ae. aegypti* abundance and the built environment, the equity-relevant associations should be interpreted as describing the joint structural context of low introgression rather than as isolating an independent causal effect of socioeconomic status. The 2010 census was used as it is the last Brazilian census with complete data; preliminary results from the 2022 census are available and socioeconomic metrics in Rio de Janeiro are very highly correlated between the two datasets. The mediation analysis (**Figure S4**) indicates SES effects are only partly explained by mosquito abundance. Additionally, release-batch quality (e.g., longevity, mating competitiveness) was not systematically recorded and represents could contribute to heterogeneity in establishment.

Finally, we estimated mosquito abundance for each trap at different calendar times, based on when initial releases and trapping began. Some of the observed differences in abundance may reflect seasonal fluctuations in mosquito populations. However, we calculated the ratio of the initial release dose to the existing mosquito population at the same calendar time, enabling us to assess intervention dosing regardless of the season.

## Conclusions

Despite many obstacles, WMP was able to implement the *w*Mel intervention in Rio de Janeiro by working with communities and local authorities throughout the study area, including less accessible, informal settlement areas. However, we found cumulative releases of *w*Mel varied significantly by area and by inhabitant and there was heterogeneity in introgression throughout the study area. In more recent release programs in Brazil, WMP has determined release doses using human population density rather than area. In Belo Horizonte, as part of the EVITA Dengue Trial, they aimed to release 88-100 *w*Mel *Ae. aegypti* per person in the first 16 weeks, targeting more homogenous introgression (27). The areas with the most successful introgression in Rio de Janeiro were higher in the socioeconomic status and achieved higher introgression with less *w*Mel released by area, highlighting inequities in the intervention outcome. After more formally testing with spatiotemporal models, we identified that lower socioeconomic status, increased built environment, and higher *Ae. aegypti* abundance were associated with lower introgression. Furthermore, we found that the ratio of mosquitoes released to baseline mosquito abundance strongly correlated with mean introgression levels, suggesting its potential utility as a planning metric. Accounting for existing *Ae. aegypti* populations when planning releases will play a key role in ensuring successful, equitable, and sustainable *Wolbachia* interventions.

## Data Availability

The minimum data set (de-identified) and accompanying code is available at the ko-laboratory/wmel-rj repository on GitHub via https://github.com/ko-laboratory/wmel-rj.

## Authors’ contributions

Conceptualization: AIK, DATC, HS, LAM, EN, PMC

Methodology: AIK, DATC, HS, LAM, EN, PMC, GRdS, ID

Investigation: EN, PMC, GRdS, GSR, TRdC

Visualization: AIK, DATC, EN, PMC, GRdS

Funding acquisition: AIK, DATC, LAM, EN

Supervision: AIK, DATC, HS, LAM

Writing – original draft: EN, PMC

Writing – review & editing: AIK, DATC, HS, LAM, EN, PMC, GRdS, ID, GSR, TRdC

## Data and materials availability

All code and de-identified data used in these analyses are available at https://github.com/ko-laboratory/wmel-rj.

## List of supplementary materials

Materials and methods

Figures S1 to S16

Tables S1 to S4

## Supplementary Materials

### Materials and Methods

#### Setting

Rio de Janeiro, the second largest city in Brazil, is a tropical city located in the Southeastern state of Rio de Janeiro (28). Covering 1,255km^2^, with a population of almost 6.7 million inhabitants, the city is characterized by varying urban landscapes. Elevation within the city ranges from 0m to 1,022m above sea level, with poorer areas generally located atop the steep, less accessible mountains (29). Temperatures range from around 21℃ in the winter/dry season to around 30℃ in the summer/wet season. Precipitation ranges from an average of 5cm in the winter/dry season to an average of 17cm in the summer/wet season.

#### Releases

World Mosquito Program Brazil performed releases of *w*Mel *Ae. aegypti* mosquitoes from August 2017 through March 2020 and in five zones of Rio de Janeiro, covering a total area of 86.8 km^2^ and a population of roughly 890,000. Releases were performed in three phases: an initial establishment phase of weekly releases for a period of 8-16 weeks, followed by up to two consolidation phases of weekly to biweekly releases performed until *w*Mel prevalence reached >60% in the area or the end of the intervention period. Approximately 350 adult *w*Mel *Ae. aegypti* mosquitoes were released at 50m intervals throughout release areas. Where possible, releases occurred using vehicles, but in less accessible areas, releases were performed on foot. Regulatory approval for field releases was obtained from the National Research Ethics Committee (CONEP, CAAE 02524513.0.1001.0008), as described in previous publications (9).

#### Trapping

Monitoring of *Ae. aegypti* abundance and *w*Mel prevalence was conducted using a network of 1,168 BG-Sentinel traps, placed at even intervals approximately every 250m and up to 16 traps per km^2^ (8,30). Traps were collected biweekly. All mosquitoes captured were morphologically identified to the species level and a random sample of up to 10 *Ae. aegypti* were tested individually for *w*Mel using quantitative PCR.

#### Covariates

We compiled data on meteorological, socioeconomic/demographic, and environmental factors potentially influencing *w*Mel introgression in the study area and time period from several sources. Meteorologic data was collected from the Brazilian National Institute of Meteorology, which maintains one meteorological station in the study area on Ilha do Governador (31). Hourly minimum, maximum, and mean temperatures (℃) and mean daily precipitation (cm) from the Galeão weather station were aggregated by week. Socioeconomic data was collected from the 2010 Brazilian Census, including several indicators of income, electricity, literacy, waste management, and piped water access at the census-tract level (32). Population density was collected from WorldPop, which uses a random forest regression tree-based mapping approach to integrate census data with other spatial datasets related to human populations to provide finer population density estimates, at 100×100m scale (33). Elevation (m) and impervious surface (%) were collected at the 30 x 30m scale from LandSat, NASA and the U.S. Geological Survey’s satellite imagery database (34). Urban Green Space data at the 10×10m scale was collected from Google Earth Engine’s Sentinel-2 database (35).

#### Descriptive Analysis

To understand the spatiotemporal heterogeneities in the release, trapping, and covariate data, as well as to assess differences in cumulative *w*Mel releases by area, we divided the study area into 11,140 100×100m cells. In each grid cell, we calculated the cumulative number of *w*Mel *Ae. aegypti* released by cell (area) and by person (inhabitants in each cell). We divided the cells into quartiles by the number of mosquitoes released by area and by person. We then described the distribution of the covariates in each quartile to understand trends in program roll-out. We also compared the number of mosquitoes released by area and by person with the average *w*Mel introgression in that cell.

#### Introgression Trajectory Modeling

To understand trends in *w*Mel introgression at a finer scale, we analyzed individual trap-level introgression through time. Given the zero-and one-inflated nature of the trap-level data due to variations in the number of total *Ae. aegypti* caught through time, we smoothed the trap-level introgression trajectories through time using generalized additive models weighted by the total number of *Ae. aegypti* captured at each time point. To create clusters of introgression trajectories through time, we conducted group-based multivariate trajectory modeling (GBMT) on the trap-level smoothed data using the ‘gbmt’ R package (36). We used model-based information criterion scores and interpretability of clustering, i.e. balance of number of traps in each cluster, to select the best number of clusters. We then extracted covariate data from a 100m radius around each trap and described covariate characteristics by cluster (37). We tested varying radii measures as a sensitivity analysis (**Table S3**).

#### Modeling Factors Influencing Introgression

To determine predictors of *w*Mel introgression across release areas, we constructed spatiotemporal regression models using the Integrated Nested Laplace Approximation (INLA) method using the R-INLA package (38). INLA approximates Bayesian inference and addresses the spatial and temporal autocorrelation of variables. All models used the trap-specific extrapolated *w*Mel-positive count at time *t* as the outcome variable, calculated by multiplying the trap prevalence of *w*Mel by the total *Ae. aegypti* caught, modeled with a negative binomial likelihood to account for overdispersion, with an offset of the total *Ae. aegypti* caught at time *t*. The models also included a time-varying autoregressive term of order 1 and a spatially varying autoregressive term using stochastic partial differential equations (**Figures S6 and S13**).

Prior to model-building, we restricted the possible meteorologic, socioeconomic, and environmental covariates included. We examined the correlation between covariates using a Spearman-rank correlation test (**Figure S3**). For highly correlated variables, we fit separate models using each and determined the best fit. For temporally varying covariates, we also examined the correlation between the model outcome (extrapolated *w*Mel-positive count) and the covariate at lags zero to eight weeks in the past using a cross correlation function (**Figure S2**). We only included significant lags in our models and restricted further based on significance. We determined whether to include covariates as linear or non-linear using generalized additive models of the covariate, the model outcome (extrapolated *w*Mel-positive count), and an offset (total *Ae. aegypti* caught) (**Figure S1**). Non-linear variables were included in the model using quintiles, fit with a random walk order of two in INLA.

To assess the robustness of our approach, we performed several sensitivity analyses of our models. We tried using monthly and yearly AR1 temporal autocorrelation terms, to ensure that we were not overfitting our model and taking away the temporally varying effects of our covariates (**Figures S7, S8**). We varied the radii surrounding the trap to 50m and 200m, to ensure that our results were not being driven by the spatial scale to which we aggregated our data (**Figures S9, S10**). We restricted our data to only areas which received releases from vehicles (**Figure S11**). Releases were performed using backpacks in areas with informal settlements and areas with security concerns, where release schedules often needed to be modified. To check that our results were not being driven by variability in releases, we restricted our analysis to areas which received the intervention as planned. We restricted our analyses to a 50% random subsample of space-time observations, to ensure we were not overfitting to our data (**Figure S12**). We restricted our analyses to space-time observations without ongoing *w*Mel releases, to ensure that recapturing recently released *w*Mel *Ae. aegypti* (i.e. contamination) was not biasing our results (**Figure S13**). To determine whether the effects of environmental, socioeconomic, and meteorological factors on introgression were being mediated by *Ae. aegypti* abundance, we fit our model without *Ae. aegypti* trap counts or previous *w*Mel introgression measures (**Figure S4**). Finally, to determine the effect of seasonality separately from the weather covariates, we included a monthly cyclic random walk term (**Figure S5**).

#### Baseline Abundance Measurement

To better understand the wildtype *w*Mel-negative *Ae. aegypti* population in the study areas and investigate predictors of introgression that could be measured at beginning of releases, we used the initial *w*Mel releases as a Mark-Release-Recapture experiment. Given that *w*Mel begins spreading through a wildtype population due to maternal inheritance and cytoplasmic incompatibility, we only included traps for which the first trap collection occurred 36 days or less after the first release (minimum of 9 days present in the data). This reduces the risk of mistakenly counting introgressed *w*Mel *Ae. aegypti* as newly released *w*Mel *Ae. aegypti*, which would lead to underestimating mosquito abundance in areas with better introgression. We also excluded traps that detected no *w*Mel-positive *Ae. aegypti* in the first trapping event. We included 85 traps in this analysis.

We calculated a previously validated metric, the Fisher-Ford Index, defined as N_t_ = Mnɸ^t^/m, where *M* is the number of *w*Mel *Ae. aegypti* released, *n* is the total number of *Ae. aegypti* captured the following week, *m* is the number of *w*Mel *Ae. aegypti* captured the following week, ɸ is the probability of daily survival of a *w*Mel *Ae. aegypti*, and *t* is the number of days between *w*Mel release and trap collection. We set ɸ to 0.82 based on previous studies (23). We included all mosquito releases in a 100m radius around each trap before the first collection event. We calculated the FFI and the ratio between the average number of mosquitoes released before the first trapping event and the FFI. We tested for associations between both of these metrics and mean introgression using generalized linear regressions. To assess robustness to the assumed ɸ, we repeated these regressions varying ɸ from 0.75-0.90 in increments of 0.01. To ensure that traps were not disproportionately sampling the same population leading to synthetic differences in abundances, we repeated these analyses aggregating traps by 100m cells (**Figure S16**).

To determine the sensitivity and specificity of the mosquitoes released/FFI ratio to predict mean introgression throughout the study period, we plotted a receiver operating characteristic (ROC) curve for a threshold of 20% mean introgression (median introgression for traps for which FFI could be calculated) (**Figure S15**).

All statistical analyses were conducted using R Statistical Software (version 4.4.1, R Foundation for Statistical Computing, Vienna, Austria) (39).

## Supplementary figures

**Figure S1.**
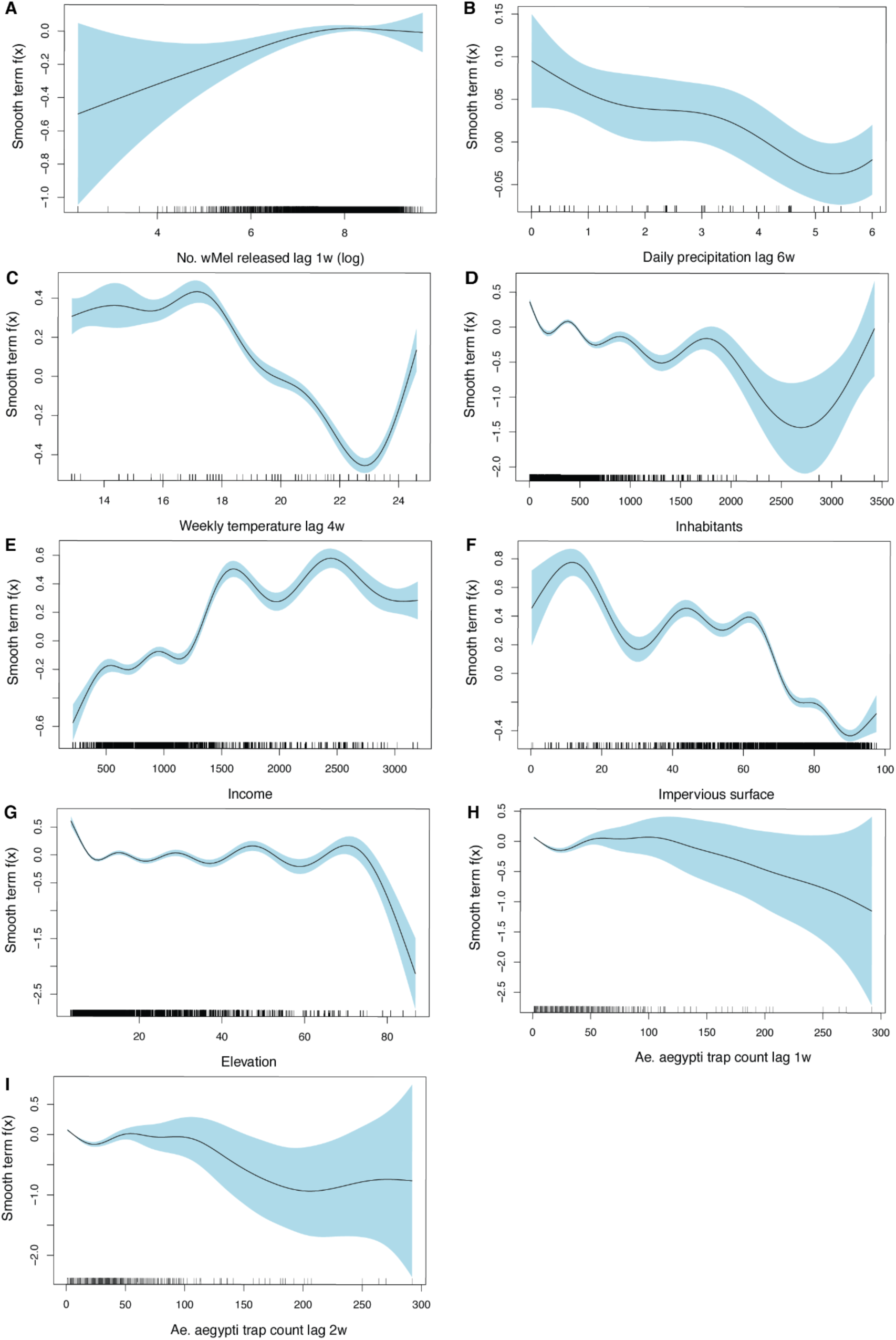
Generalized additive models used to determine covariate linearity for spatiotemporal INLA models. Univariate generalized additive models (GAMs) were run on each variable prior to inclusion in the spatiotemporal INLA models, to determine the linear of their relationship with the outcome variable of extrapolated *w*Mel count (prevalence*total *Ae. aegypti* caught by the trap). The results of the variables included in the final models are shown above. All GAMs included the outcome of extrapolated *w*Mel count, the variable of interest shown on the x-axis, and an offset of total *Ae. aegypti* trap count through time at the trap-level. The association found is shown by the black line, surrounded by a 95% confidence interval. Along the x-axis, there is a rug plot describing the distribution of the data.

**Figure S2.**
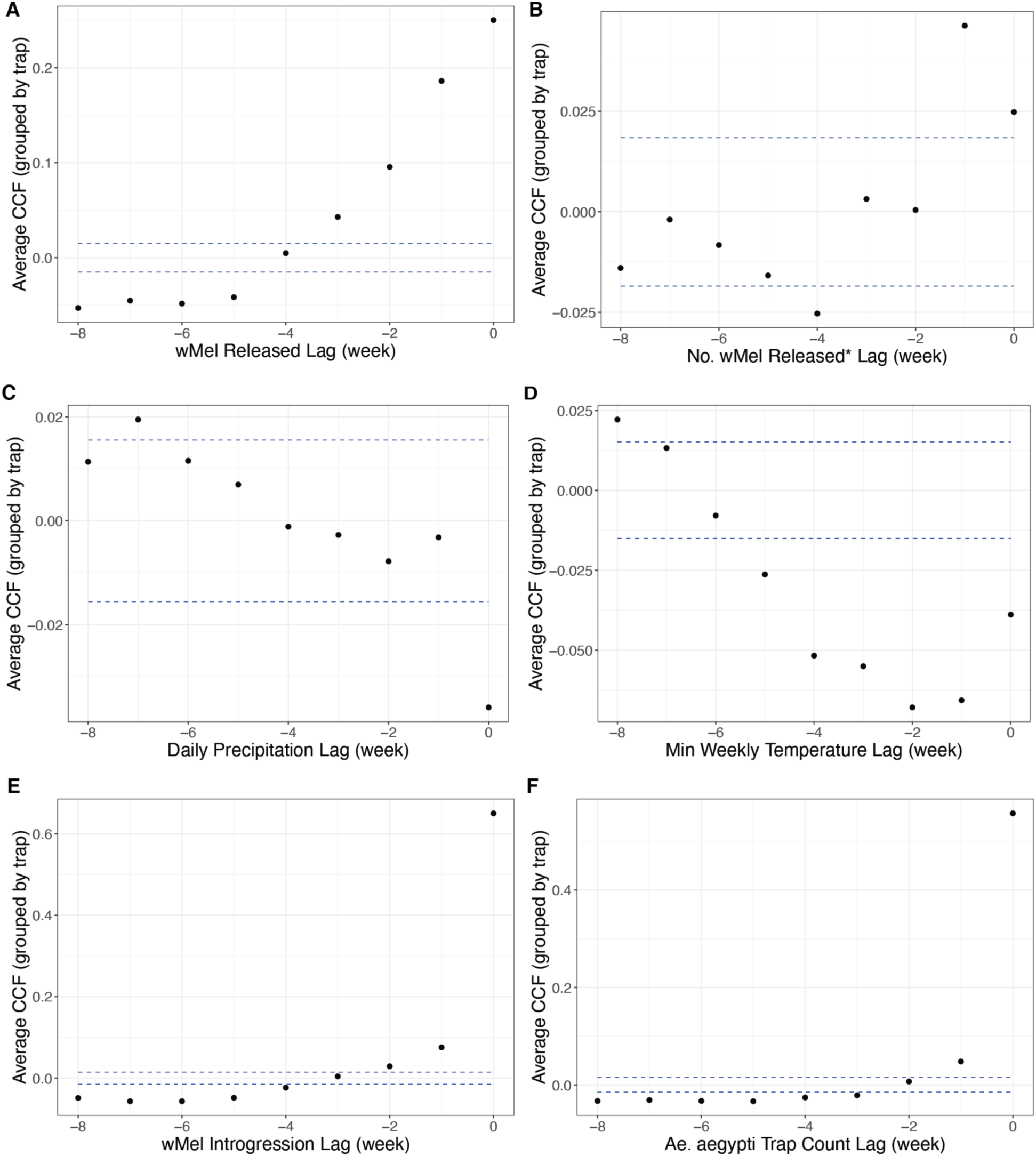
Cross correlation function results used to determine significant temporal lags of covariates to be tested in spatiotemporal INLA models. The correlation of all temporally-varying variables were tested using historical lags 0-8 weeks against the outcome variable of extrapolated *w*Mel count (prevalence*total *Ae. aegypti* caught by the trap) for each trap. The results of the variables included in the final models are shown above. The mean correlation coefficient for all traps is shown for each lag by the black dots. The blue dashed lines indicate the 95% thresholds of significance. Only lags with coefficients outside of the threshold lines prior to lags becoming nonsignificant were included in the models.

**Figure S3.**
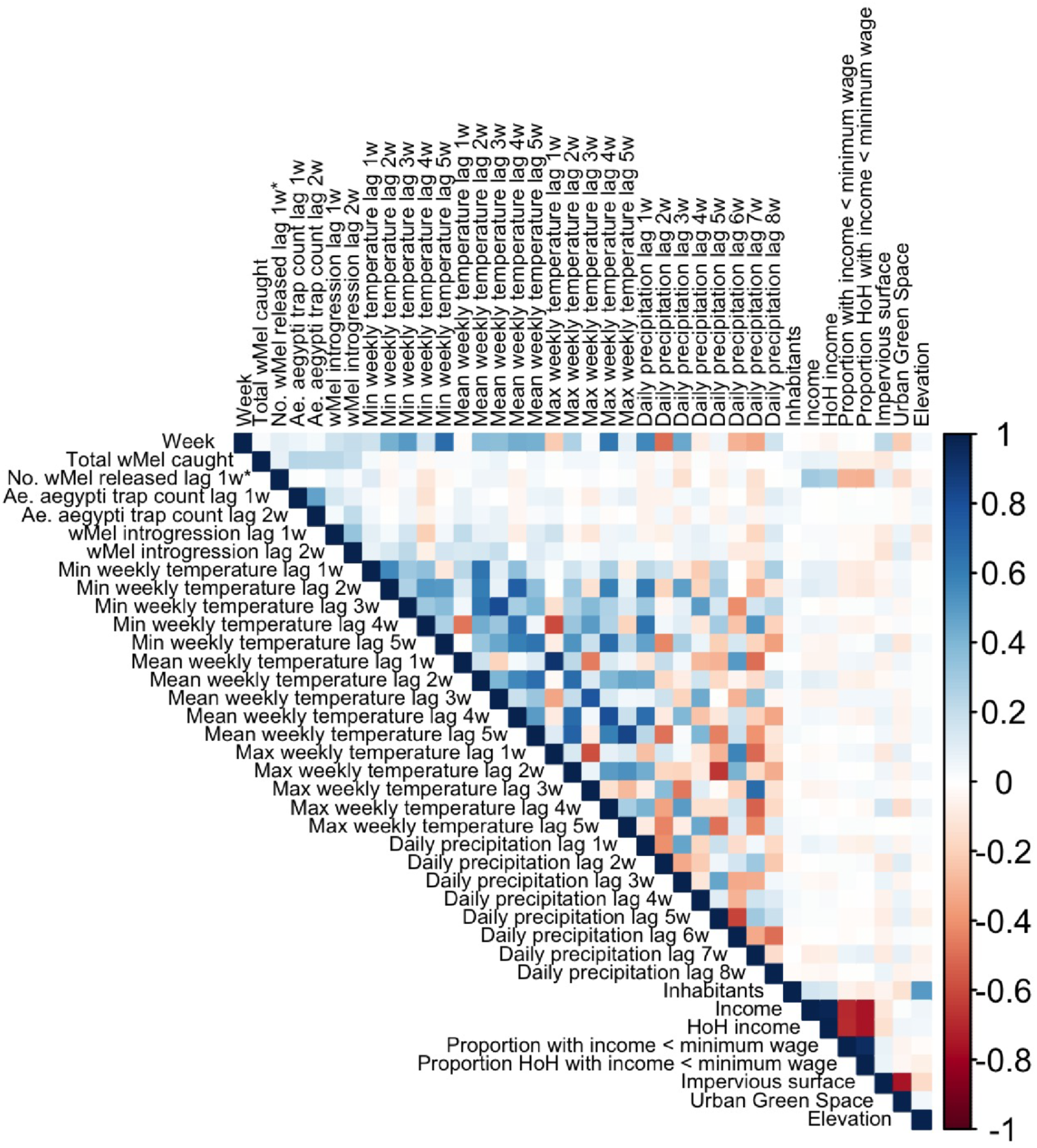
Correlation of covariates tested in spatiotemporal models. Correlation was estimated using a Spearman-rank correlation test arranged by week date and grouped by trap. Darker blue cells indicate high positive temporal correlation. White cells (0) indicate no temporal correlation. Darker red cells indicate high negative temporal correlation. *w*Mel released lag 1w could not be included due to its binary structure.

**Figure S4.**
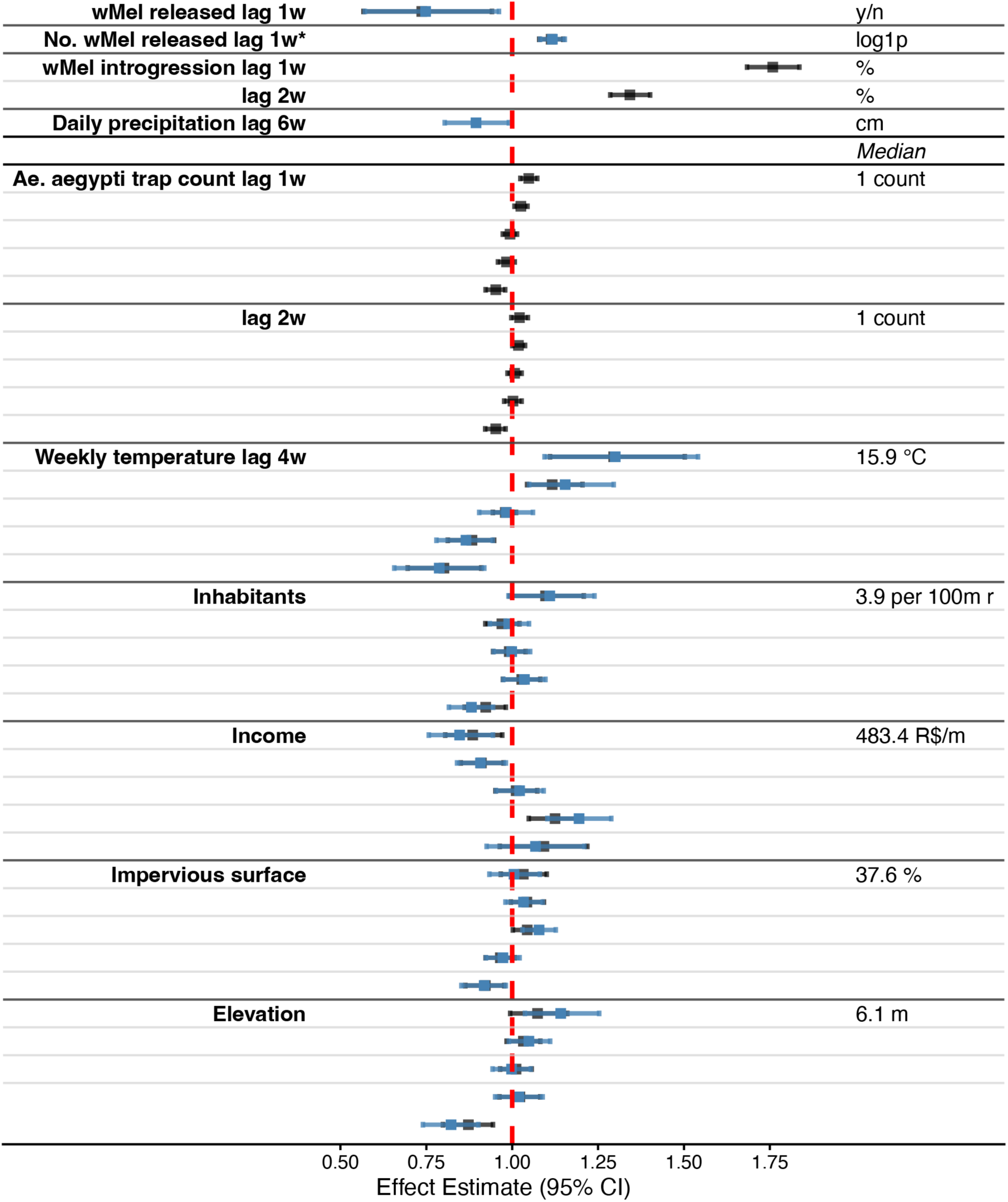
Spatiotemporal analysis of environmental, SE and meteorological factors associated without *w*Mel introgression. (Conditional R^2^=0.75). The effects of the original model are shown in black, while the sensitivity analysis model effects are shown in blue. The outcome of the model is extrapolated *w*Mel count (*w*Mel prevalence\**Ae. aegypti* caught). Non-linear effects were run as quintiles, depicted in the distribution plots, with red lines marking the boundaries, and medians and units of measurement in the right text column. All data was aggregated by week and scaled to the 100m radius surrounding traps. ‘*w*Mel released lag 1w’ is a binary term, indicating whether releases occurred; ‘No. *w*Mel released lag 1w*’ is the log(1+x) or log1p number of mosquitoes released; ‘Daily precipitation lag 6w’ is the mean daily precipitation, in centimeters; ‘Weekly temperature lag 4w’ is the minimum weekly temperature, in degrees Celsius; ‘Inhabitants’ is the population density; ‘Income’ is the mean income in R$ for persons over 10; ‘Impervious surface’ is the percent area comprised of water-impervious surfaces; ‘Elevation’ is measured in meters.

**Figure S5.**
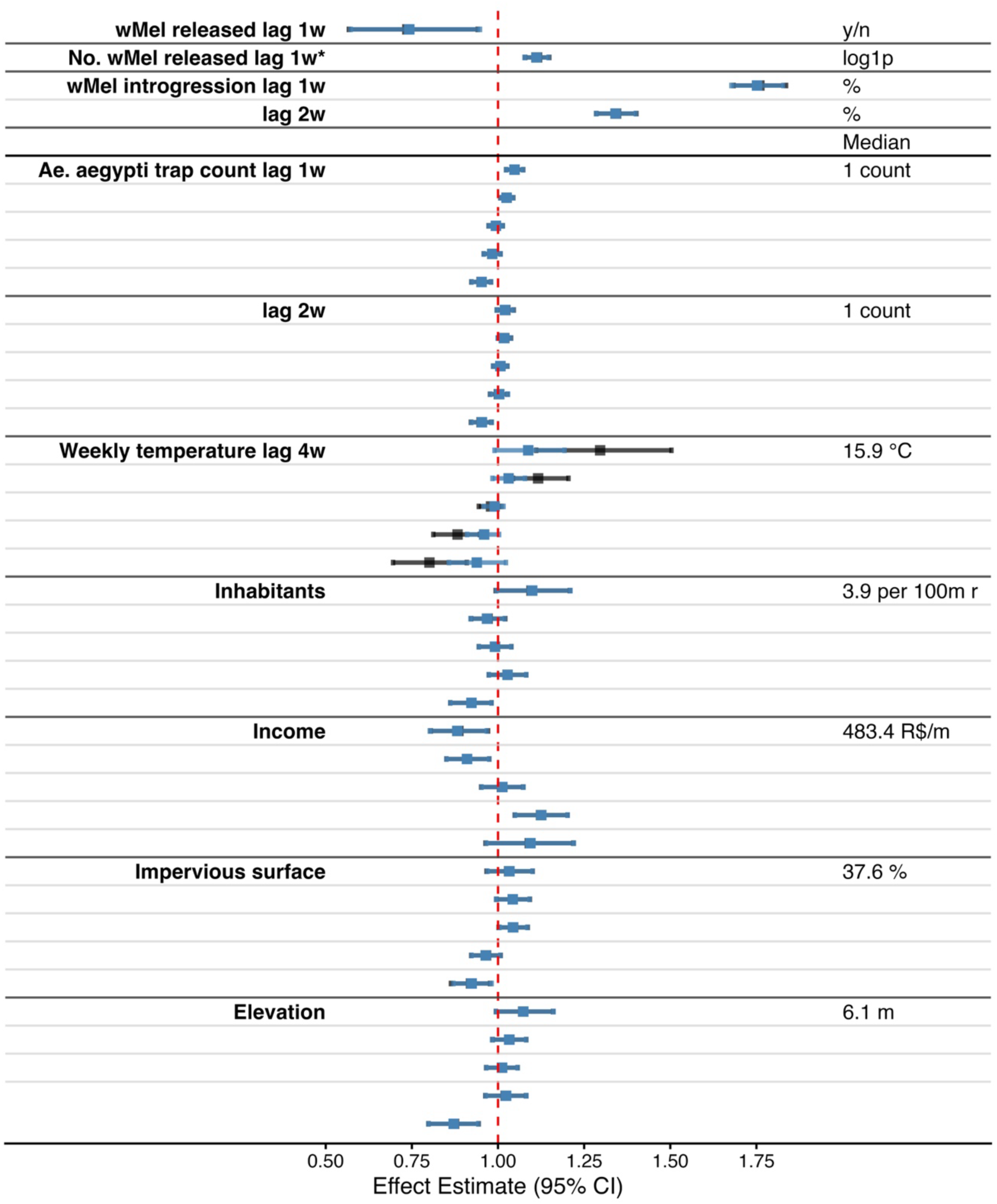
Model with monthly cyclic random walk term for seasonality: Spatiotemporal analysis of factors measured during the intervention associated with *w*Mel introgression. The effects of the original model are shown in black, while the sensitivity analysis model effects are shown in blue. Several lags of variables only show effects from the sensitivity analysis model, as they were not significant in the main model. The outcome is extrapolated *w*Mel count (*w*Mel prevalence\**Ae. aegypti* caught). Terms above *Median* were included as linear, and below as non-linear. Non-linear effects were run as quintiles, depicted in the distribution plots, with red lines marking the boundaries, and medians and units of measurement in the text column. Data was aggregated by week and scaled to the 100m radius surrounding traps. ‘*w*Mel released lag 1w’ is a binary term, indicating whether releases occurred; ‘No. *w*Mel released lag 1w*’ is the log(1+x) number of mosquitoes released; ‘*w*Mel introgression lag 1w, 2w’ is the percent *w*Mel-positive mosquitoes; ‘Daily precipitation lag 6w’ is the mean daily precipitation, in centimeters; ‘*Ae. aegypti* trap count lag 1w, 2w’ are measures of *Ae. aegypti* abundance, distribution tails not depicted; ‘Weekly temperature lag 4w’ is the minimum weekly temperature, in Celsius; ‘Inhabitants’ is the population density; ‘HoH income’ is the mean head of household income, but the main model used the original ‘Income’ variable; ‘Impervious surface’ is the percent area comprised of water-impervious surfaces; ‘Elevation’ is in meters.

**Figure S6.**
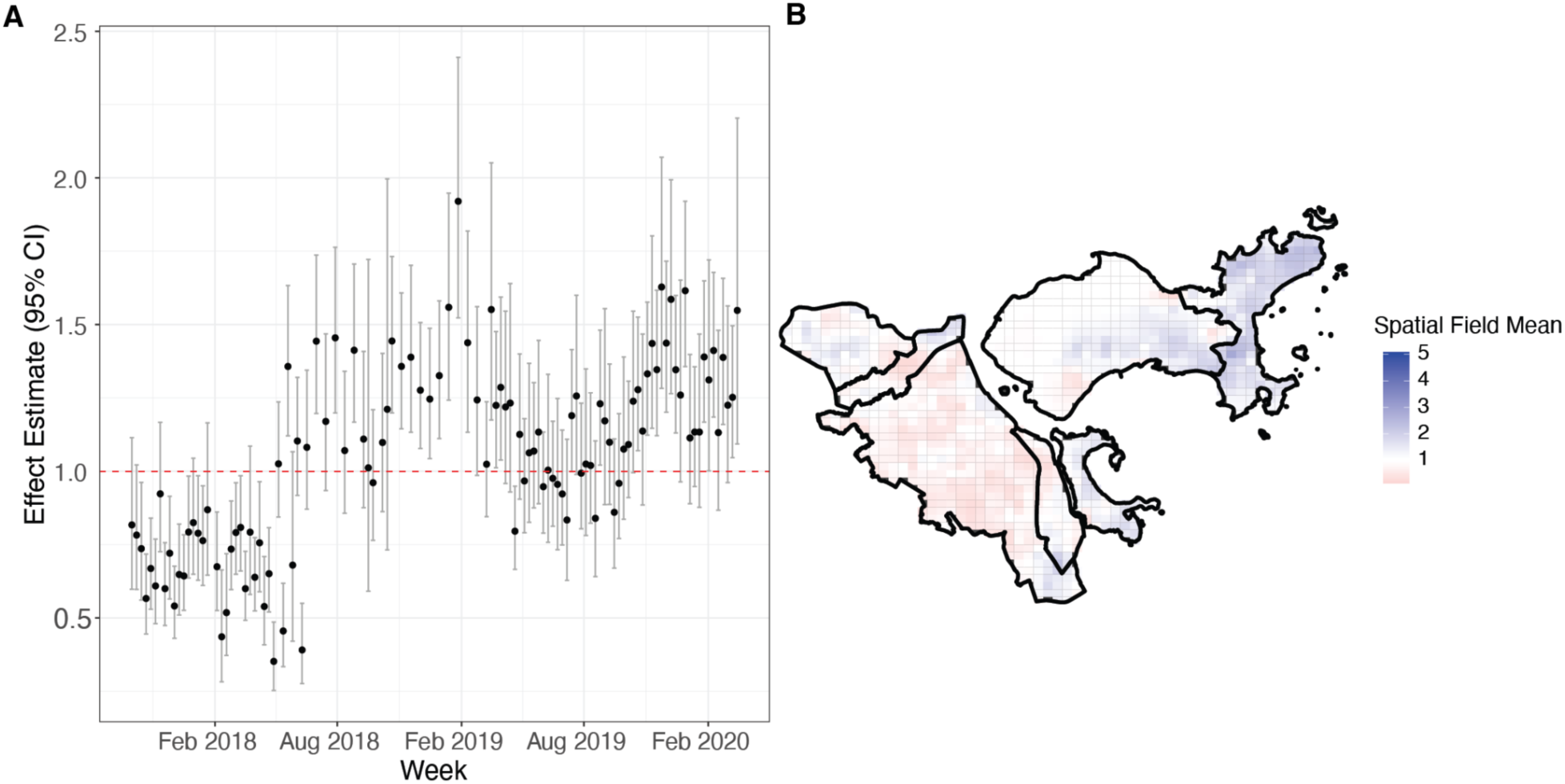
Model temporal and spatial autoregressive terms. A) The temporal autoregressive term was fit using a weekly autoregressive order one (AR1) term in INLA. The weekly autoregressive effect fit is shown by the black dots, with corresponding 95% confidence intervals in gray. The effects show slight seasonality, as evidenced by the slight peaks and troughs in effects. There is also a switch from negative to positive around week 40, which corresponds to the onset of consolidation releases in the RJ3 areas. This is likely due to the frequency-dependent fitness effects of *w*Mel introgression (cytoplasmic incompatibility and maternal inheritance). B) The spatial autoregressive term was fit using stochastic partial differentiation equations (SPDE) fit to a mesh of trap-level geolocations. The spatial field mean shows the average spatial variation across the study area through time. Areas with lower spatial field means, shown in red, likely have a weaker underlying spatial process. Whereas areas with higher spatial field means, shown in blue, have a stronger underlying spatial process and may correspond to hotspots of high introgression.

**Figure S7.**
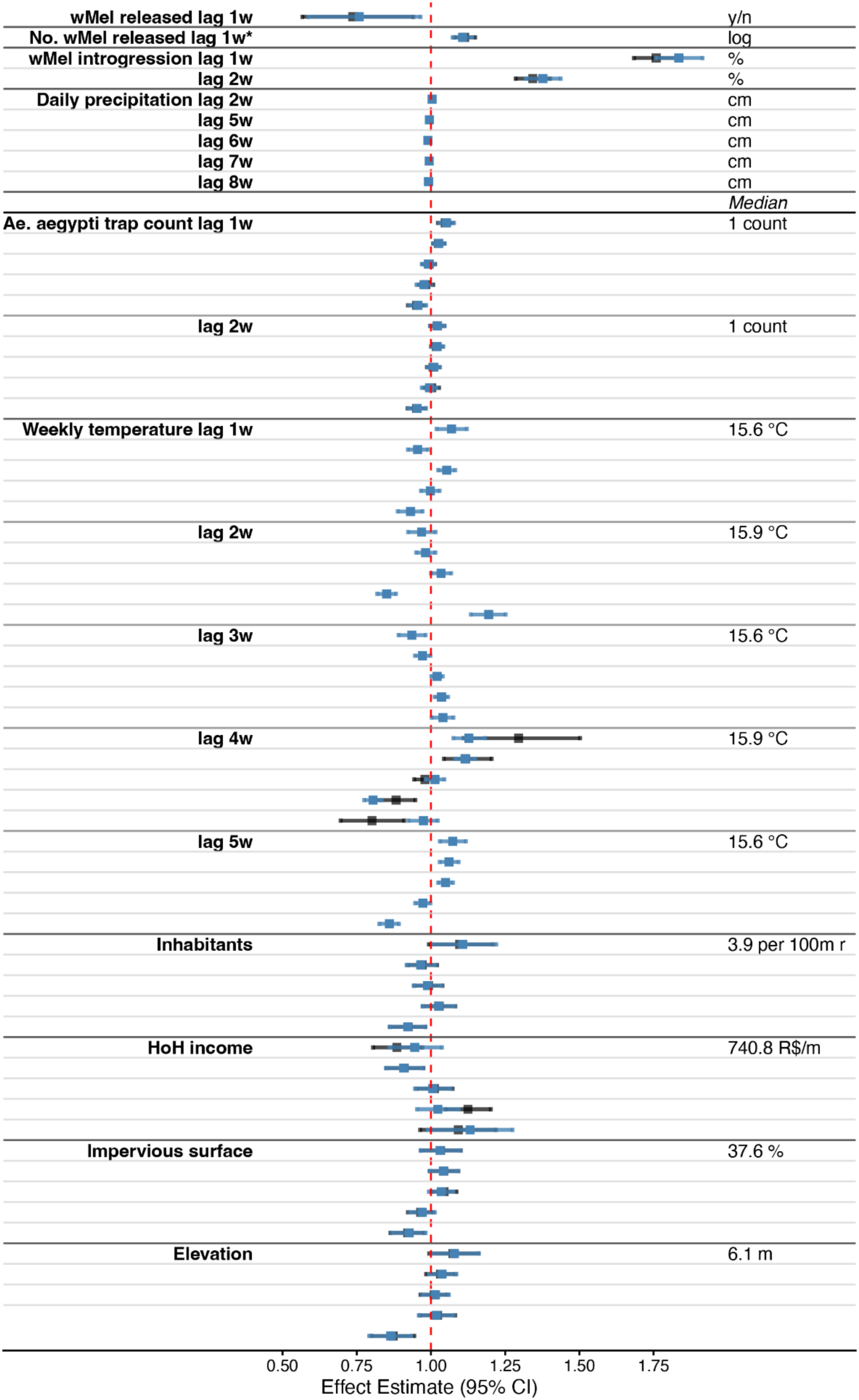
Model with monthly AR1 term: Spatiotemporal analysis of factors measured during the intervention associated with *w*Mel introgression. The effects of the original model are shown in black, while the sensitivity analysis model effects are shown in blue. Several lags of variables only show effects from the sensitivity analysis model, as they were not significant in the main model. The outcome is extrapolated *w*Mel count (*w*Mel prevalence\**Ae. aegypti* caught). Terms above *Median* were included as linear, and below as non-linear. Non-linear effects were run as quintiles, depicted in the distribution plots, with red lines marking the boundaries, and medians and units of measurement in the text column. Data was aggregated by week and scaled to the 100m radius surrounding traps. ‘*w*Mel released lag 1w’ is a binary term, indicating whether releases occurred; ‘No. *w*Mel released lag 1w*’ is the log(1+x) number of mosquitoes released; ‘*w*Mel introgression lag 1w, 2w’ is the percent *w*Mel-positive mosquitoes; ‘Daily precipitation lag 6w’ is the mean daily precipitation, in centimeters; ‘*Ae. aegypti* trap count lag 1w, 2w’ are measures of *Ae. aegypti* abundance, distribution tails not depicted; ‘Weekly temperature lag 4w’ is the minimum weekly temperature, in Celsius; ‘Inhabitants’ is the population density; ‘HoH income’ is the mean head of household income, but the main model used the original ‘Income’ variable; ‘Impervious surface’ is the percent area comprised of water-impervious surfaces; ‘Elevation’ is in meters.

**Figure S8.**
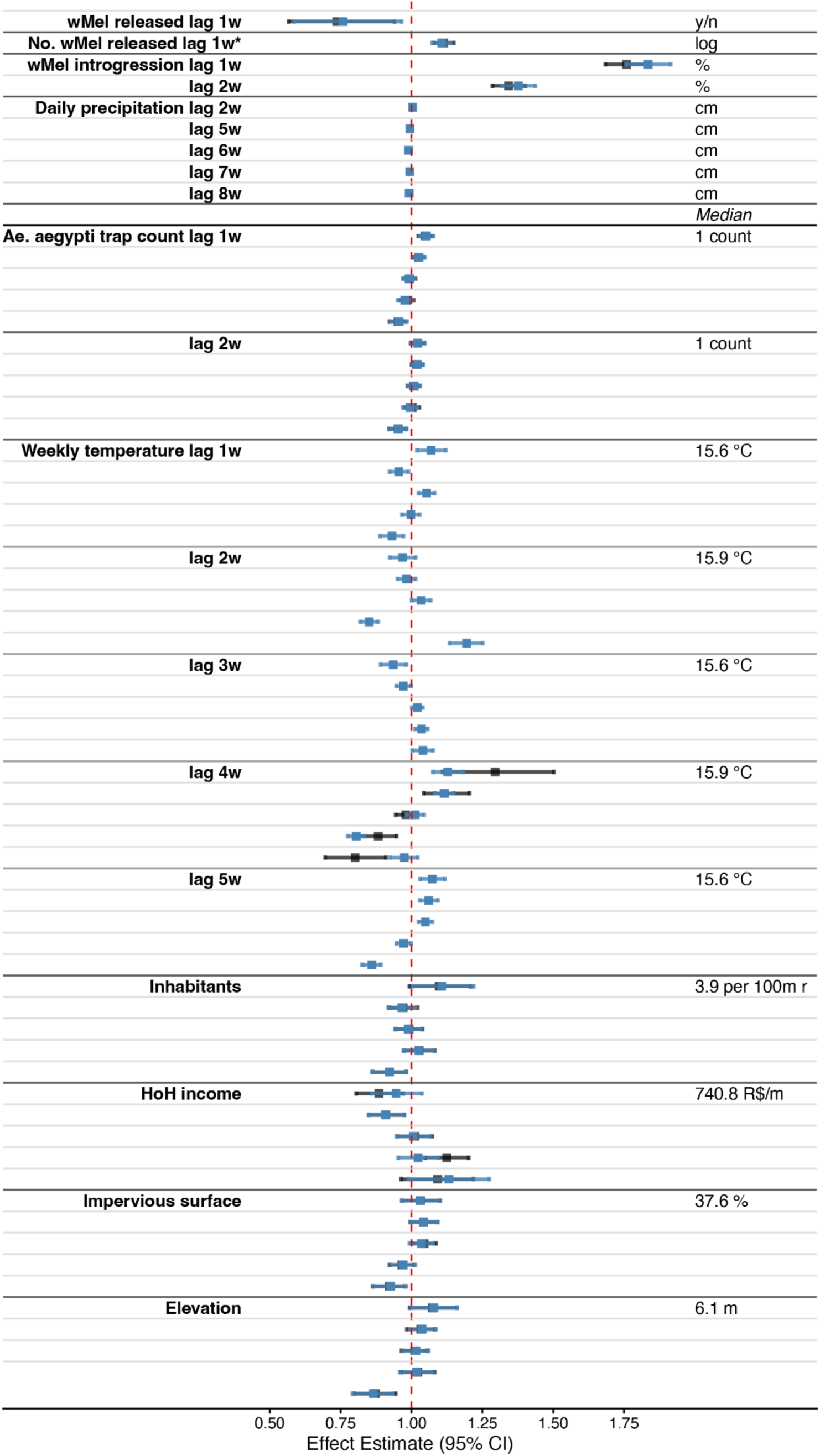
Model with yearly AR1 term: Spatiotemporal analysis of factors measured during the intervention associated with *w*Mel introgression. The effects of the original model are shown in black, while the sensitivity analysis model effects are shown in blue. Several lags of variables only show effects from the sensitivity analysis model, as they were not significant in the main model. The outcome is extrapolated *w*Mel count (*w*Mel prevalence\**Ae. aegypti* caught). Terms above *Median* were included as linear, and below as non-linear. Non-linear effects were run as quintiles, depicted in the distribution plots, with red lines marking the boundaries, and medians and units of measurement in the text column. Data was aggregated by week and scaled to the 100m radius surrounding traps. ‘*w*Mel released lag 1w’ is a binary term, indicating whether releases occurred; ‘No. *w*Mel released lag 1w*’ is the log(1+x) number of mosquitoes released; ‘*w*Mel introgression lag 1w, 2w’ is the percent *w*Mel-positive mosquitoes; ‘Daily precipitation lag 6w’ is the mean daily precipitation, in centimeters; ‘*Ae. aegypti* trap count lag 1w, 2w’ are measures of *Ae. aegypti* abundance, distribution tails not depicted; ‘Weekly temperature lag 4w’ is the minimum weekly temperature, in Celsius; ‘Inhabitants’ is the population density; ‘HoH income’ is the mean head of household income, but the main model used the original ‘Income’ variable; ‘Impervious surface’ is the percent area comprised of water-impervious surfaces; ‘Elevation’ is in meters.

**Figure S9.**
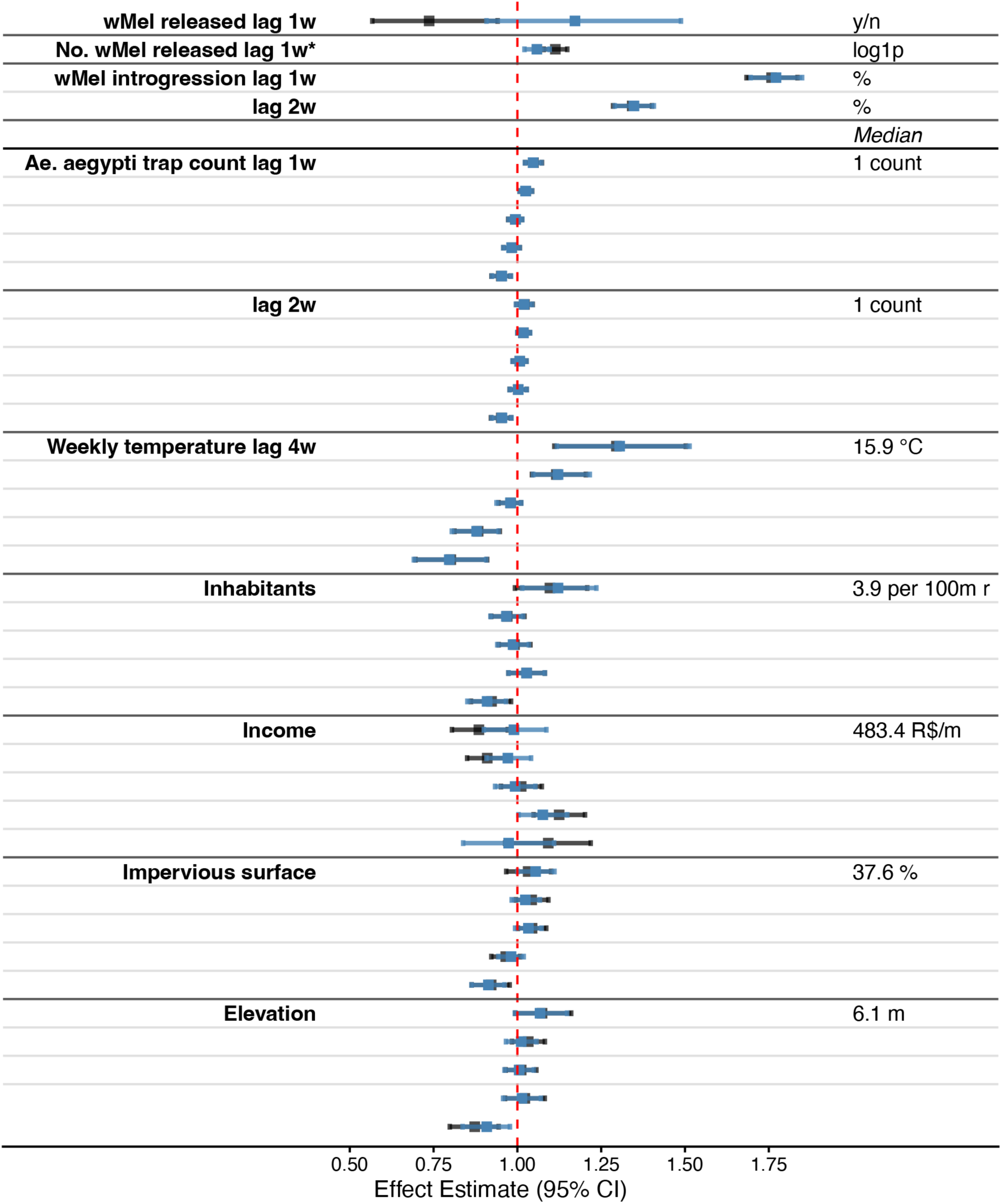
Model using covariates from a 50m radius surrounding traps: Spatiotemporal analysis of factors measured during the intervention associated with *w*Mel introgression. The effects of the original model are shown in black, while the sensitivity analysis model effects are shown in blue. The outcome is extrapolated *w*Mel count (*w*Mel prevalence\**Ae. aegypti* caught). Terms above *Median* were included as linear, and below as non-linear. Non-linear effects were run as quintiles, depicted in the distribution plots, with red lines marking the boundaries, and medians and units of measurement in the text column. Data was aggregated by week and scaled to the 50m radius surrounding traps. ‘*w*Mel released lag 1w’ is a binary term, indicating whether releases occurred; ‘No. *w*Mel released lag 1w*’ is the log(1+x) number of mosquitoes released; ‘*w*Mel introgression lag 1w, 2w’ is the percent *w*Mel-positive mosquitoes; ‘*Ae. aegypti* trap count lag 1w, 2w’ are measures of *Ae. aegypti* abundance, distribution tails not depicted; ‘Weekly temperature lag 4w’ is the minimum weekly temperature, in Celsius; ‘Inhabitants’ is the population density; ‘Income’ is the mean income in R$ for persons over 10; ‘Impervious surface’ is the percent area comprised of water-impervious surfaces; ‘Elevation’ is in meters.

**Figure S10.**
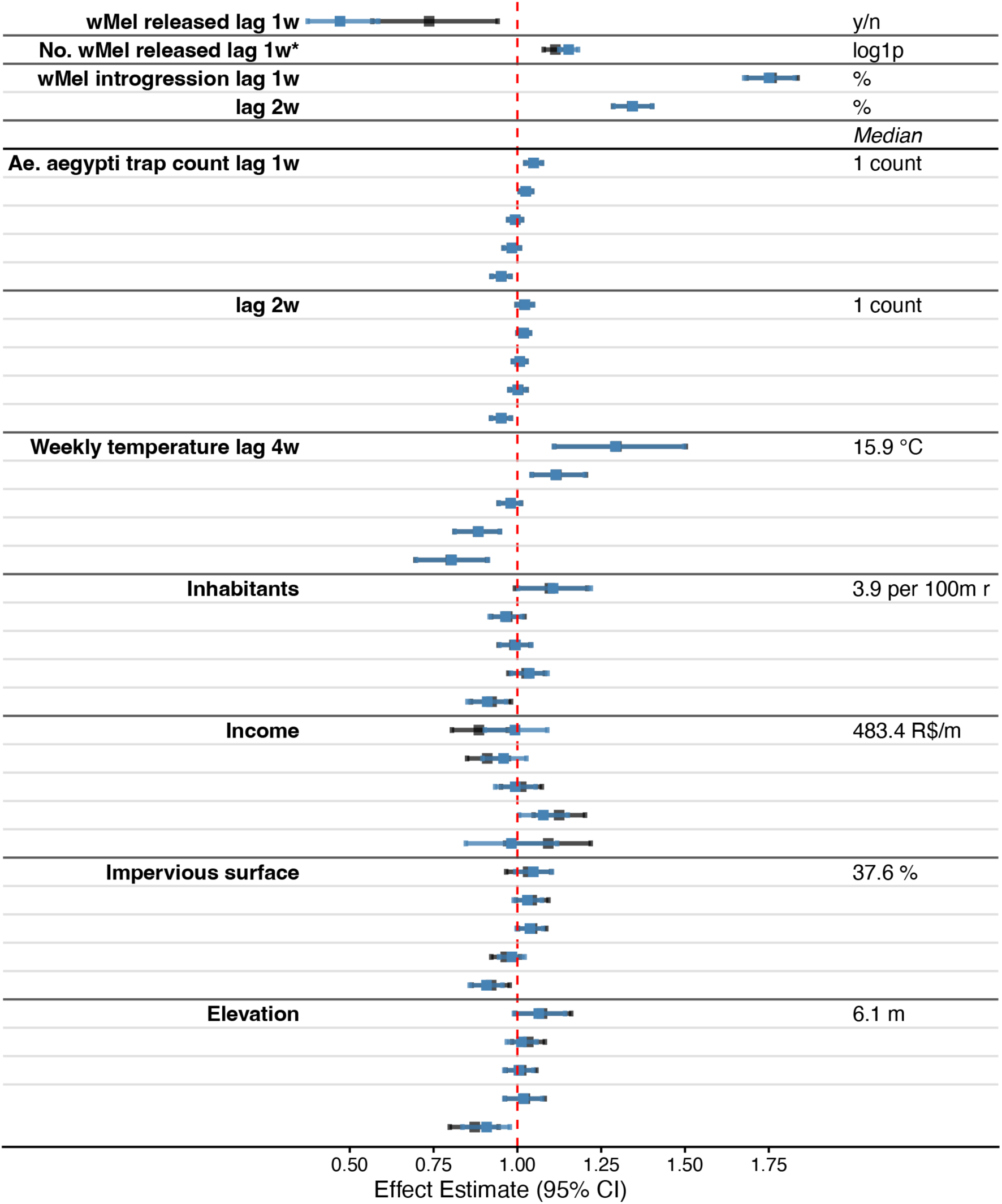
Model using covariates from a 200m radius surrounding traps: Spatiotemporal analysis of factors measured during the intervention associated with *w*Mel introgression. The effects of the original model are shown in black, while the sensitivity analysis model effects are shown in blue. The outcome is extrapolated *w*Mel count (*w*Mel prevalence\**Ae. aegypti* caught). Terms above *Median* were included as linear, and below as non-linear. Non-linear effects were run as quintiles, depicted in the distribution plots, with red lines marking the boundaries, and medians and units of measurement in the text column. Data was aggregated by week and scaled to the 200m radius surrounding traps. ‘*w*Mel released lag 1w’ is a binary term, indicating whether releases occurred; ‘No. *w*Mel released lag 1w*’ is the log(1+x) number of mosquitoes released; ‘*w*Mel introgression lag 1w, 2w’ is the percent *w*Mel-positive mosquitoes; ‘*Ae. aegypti* trap count lag 1w, 2w’ are measures of *Ae. aegypti* abundance, distribution tails not depicted; ‘Weekly temperature lag 4w’ is the minimum weekly temperature, in Celsius; ‘Inhabitants’ is the population density; ‘Income’ is the mean income in R$ for persons over 10; ‘Impervious surface’ is the percent area comprised of water-impervious surfaces; ‘Elevation’ is in meters.

**Figure S11.**
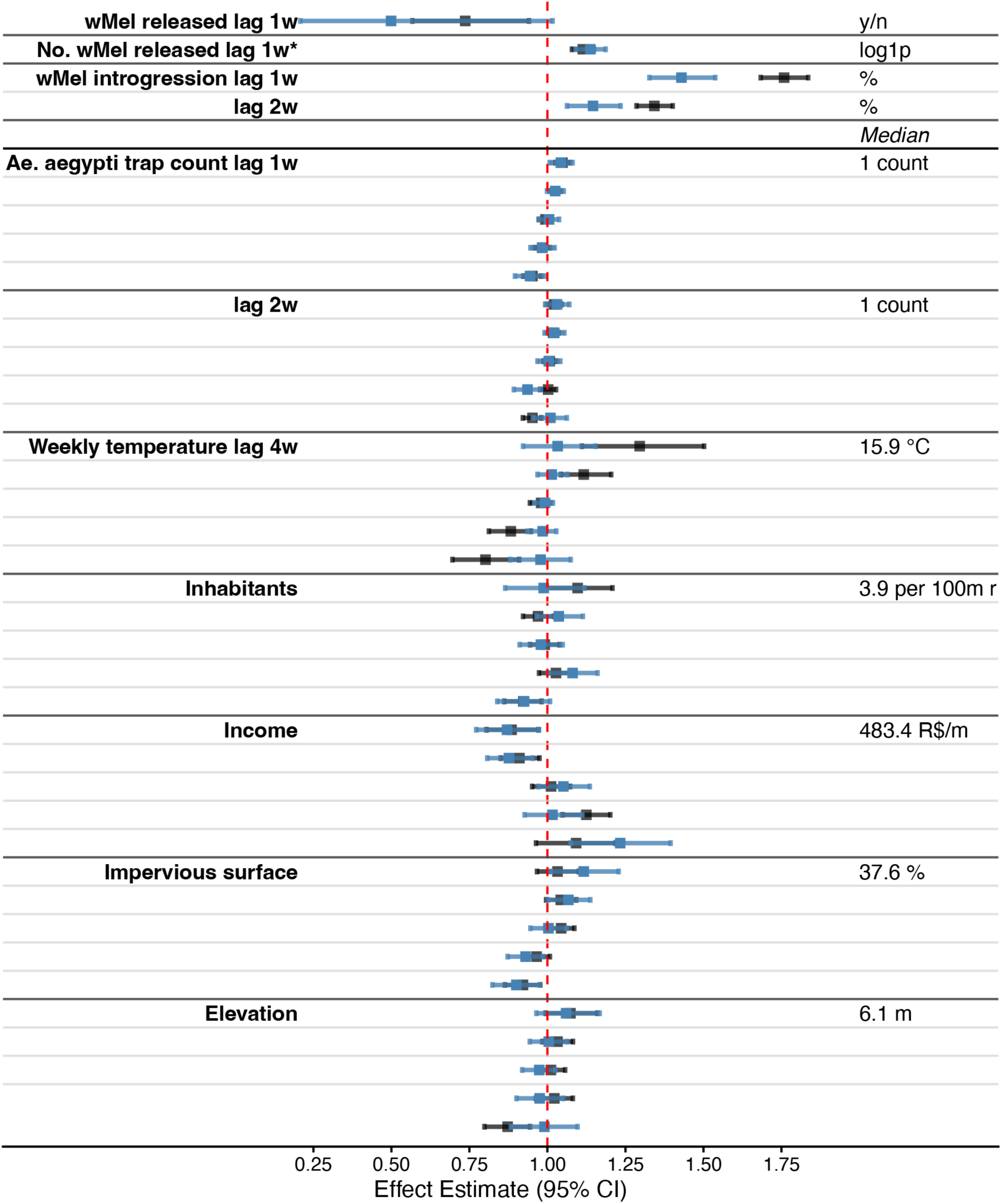
Model using only space-time observations from areas with vehicle-based *w*Mel releases: Spatiotemporal analysis of factors measured during the intervention associated with *w*Mel introgression. The effects of the original model are shown in black, while the sensitivity analysis model effects are shown in blue. The outcome is extrapolated *w*Mel count (*w*Mel prevalence\**Ae. aegypti* caught). Terms above *Median* were included as linear, and below as non-linear. Non-linear effects were run as quintiles, depicted in the distribution plots, with red lines marking the boundaries, and medians and units of measurement in the text column. Data was aggregated by week and scaled to the 100m radius surrounding traps. ‘*w*Mel released lag 1w’ is a binary term, indicating whether releases occurred; ‘No. *w*Mel released lag 1w*’ is the log(1+x) number of mosquitoes released; ‘*w*Mel introgression lag 1w, 2w’ is the percent *w*Mel-positive mosquitoes; ‘*Ae. aegypti* trap count lag 1w, 2w’ are measures of *Ae. aegypti* abundance, distribution tails not depicted; ‘Weekly temperature lag 4w’ is the minimum weekly temperature, in Celsius; ‘Inhabitants’ is the population density; ‘Income’ is the mean income in R$ for persons over 10; ‘Impervious surface’ is the percent area comprised of water-impervious surfaces; ‘Elevation’ is in meters.

**Figure S12.**
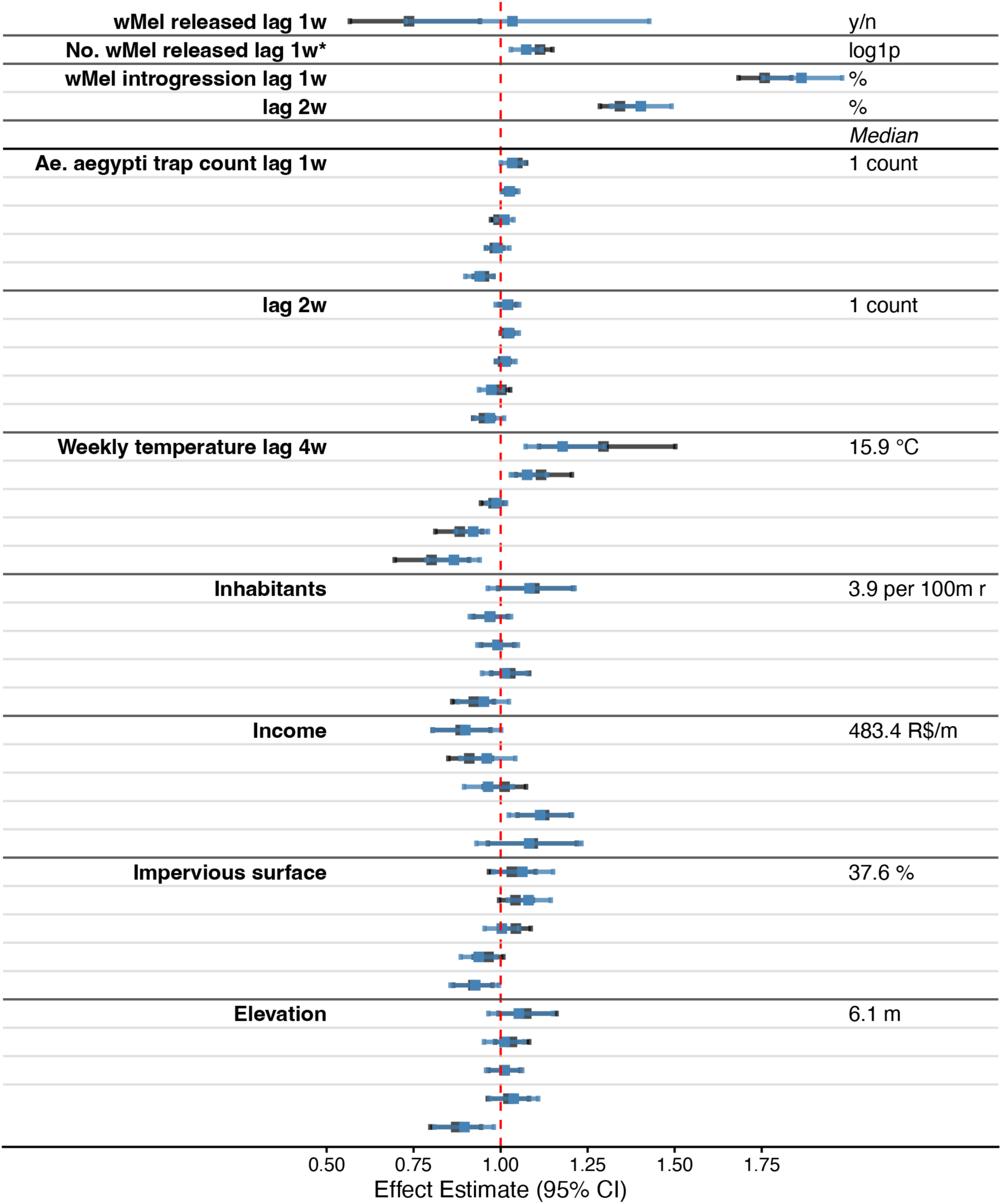
Model using a random 50% subsample of space-time observations: Spatiotemporal analysis of factors measured during the intervention associated with *w*Mel introgression. The effects of the original model are shown in black, while the sensitivity analysis model effects are shown in blue. The outcome is extrapolated *w*Mel count (*w*Mel prevalence\**Ae. aegypti* caught). Terms above *Median* were included as linear, and below as non-linear. Non-linear effects were run as quintiles, depicted in the distribution plots, with red lines marking the boundaries, and medians and units of measurement in the text column. Data was aggregated by week and scaled to the 100m radius surrounding traps. ‘*w*Mel released lag 1w’ is a binary term, indicating whether releases occurred; ‘No. *w*Mel released lag 1w*’ is the log number of mosquitoes released; ‘*w*Mel introgression lag 1w, 2w’ is the percent *w*Mel-positive mosquitoes; ‘*Ae. aegypti* trap count lag 1w, 2w’ are measures of *Ae. aegypti* abundance, distribution tails not depicted; ‘Weekly temperature lag 4w’ is the minimum weekly temperature, in Celsius; ‘Inhabitants’ is the population density; ‘Income’ is the mean income in R$ for persons over 10; ‘Impervious surface’ is the percent area comprised of water-impervious surfaces; ‘Elevation’ is in meters.

**Figure S13.**
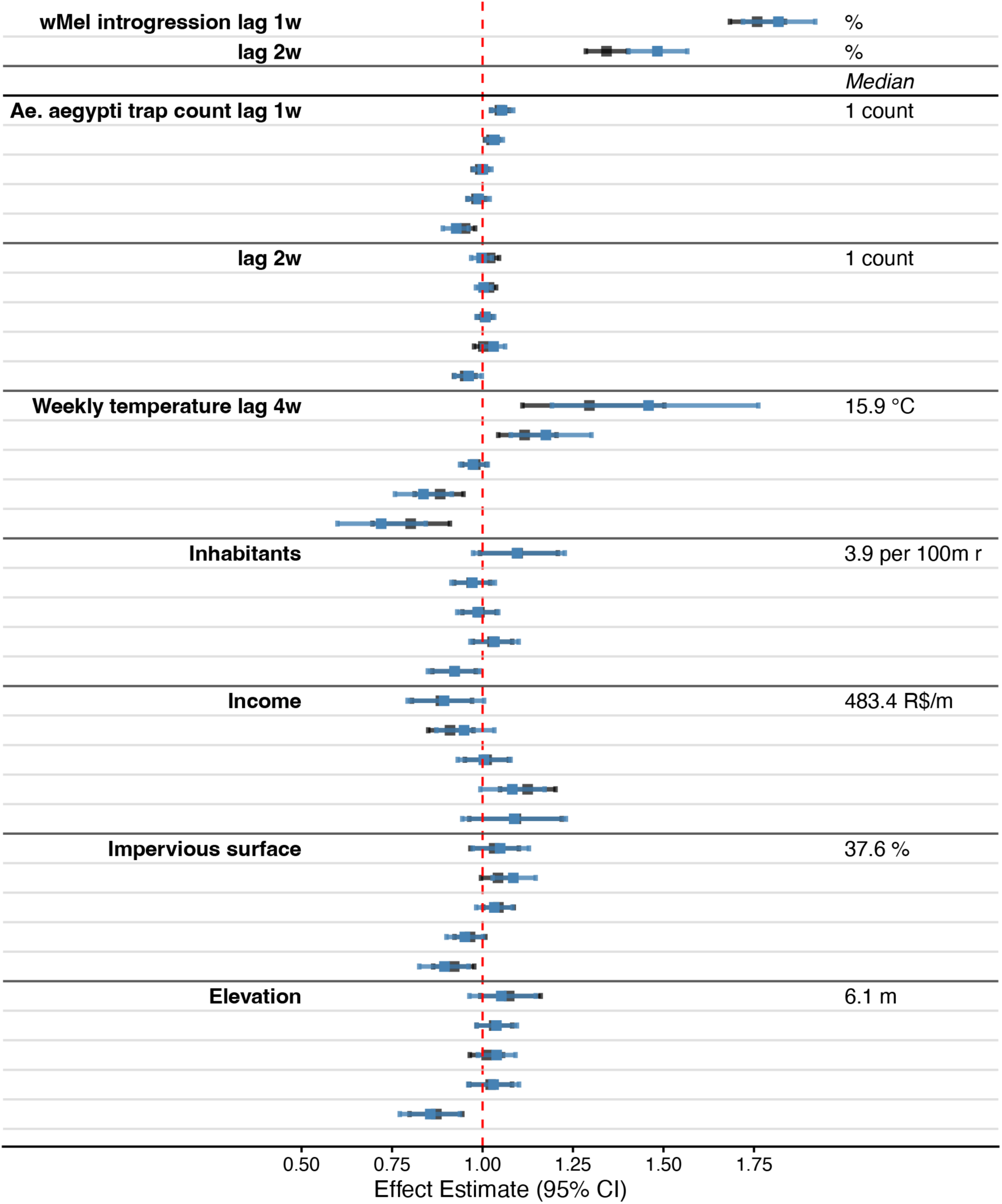
Model using only space-time observations from non-release periods: Spatiotemporal analysis of factors measured during the intervention associated with *w*Mel introgression. The effects of the original model are shown in black, while the sensitivity analysis model effects are shown in blue. The outcome is extrapolated *w*Mel count (*w*Mel prevalence\**Ae. aegypti* caught). Terms above *Median* were included as linear, and below as non-linear. Non-linear effects were run as quintiles, depicted in the distribution plots, with red lines marking the boundaries, and medians and units of measurement in the text column. Data was aggregated by week and scaled to the 100m radius surrounding traps. ‘*w*Mel introgression lag 1w, 2w’ is the percent *w*Mel-positive mosquitoes; ‘*Ae. aegypti* trap count lag 1w, 2w’ are measures of *Ae. aegypti* abundance, distribution tails not depicted; ‘Weekly temperature lag 4w’ is the minimum weekly temperature, in Celsius; ‘Inhabitants’ is the population density; ‘Income’ is the mean income in R$ for persons over 10; ‘Impervious surface’ is the percent area comprised of water-impervious surfaces; ‘Elevation’ is in meters.

**Figure S14.**
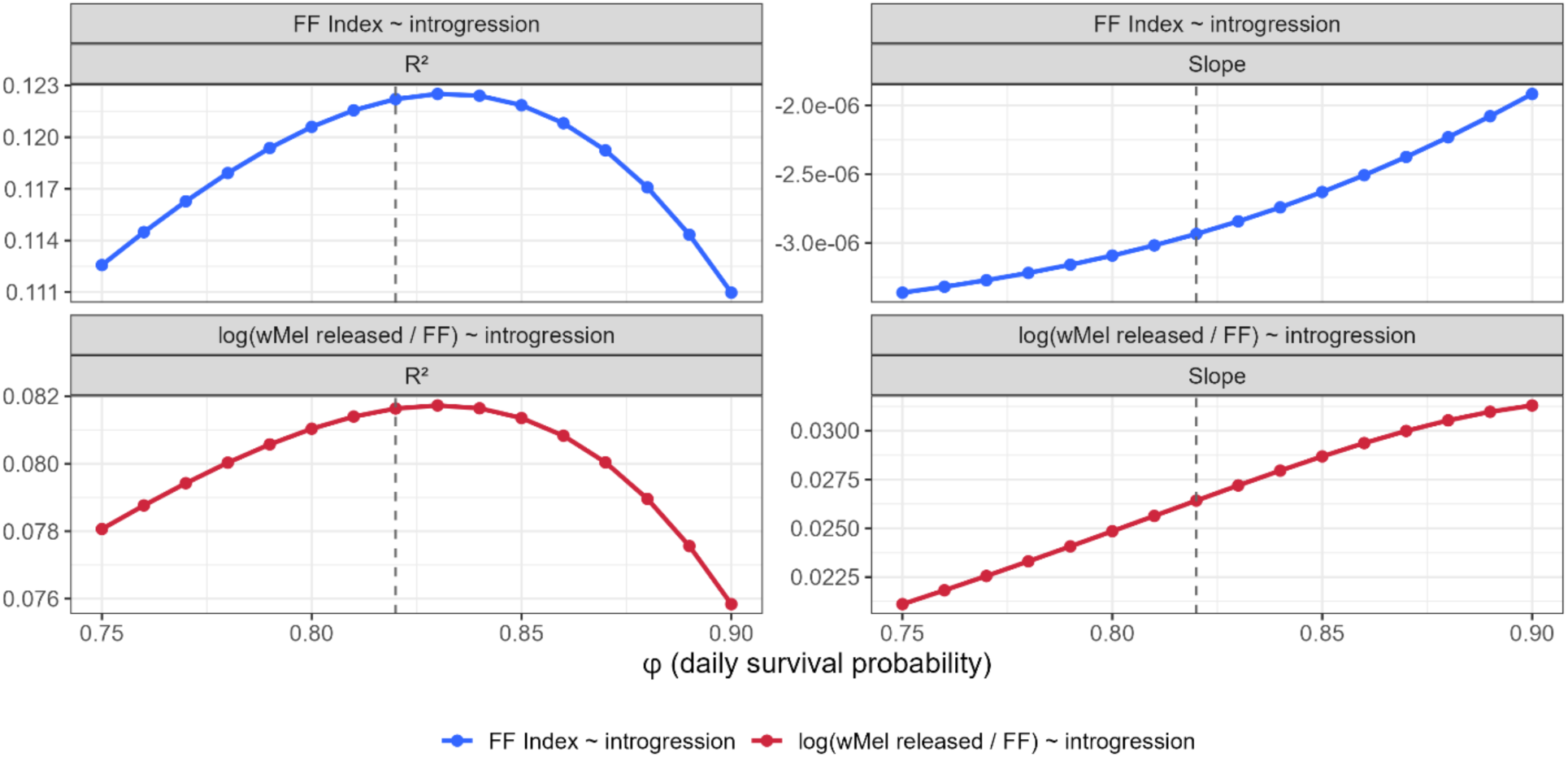
**Sensitivity of associations between Fisher-Ford Index and *w*Mel introgression to daily survival probability assumptions**. Facets on the left show explained deviance (R^2^) and facets on the right show slope for linear regression of Fisher-Ford (FF) Index and ratio of *w*Mel released (mean of releases before first trapping event) to Fisher-Ford Index versus mean *w*Mel introgression (log-transformed).

**Figure S15.**
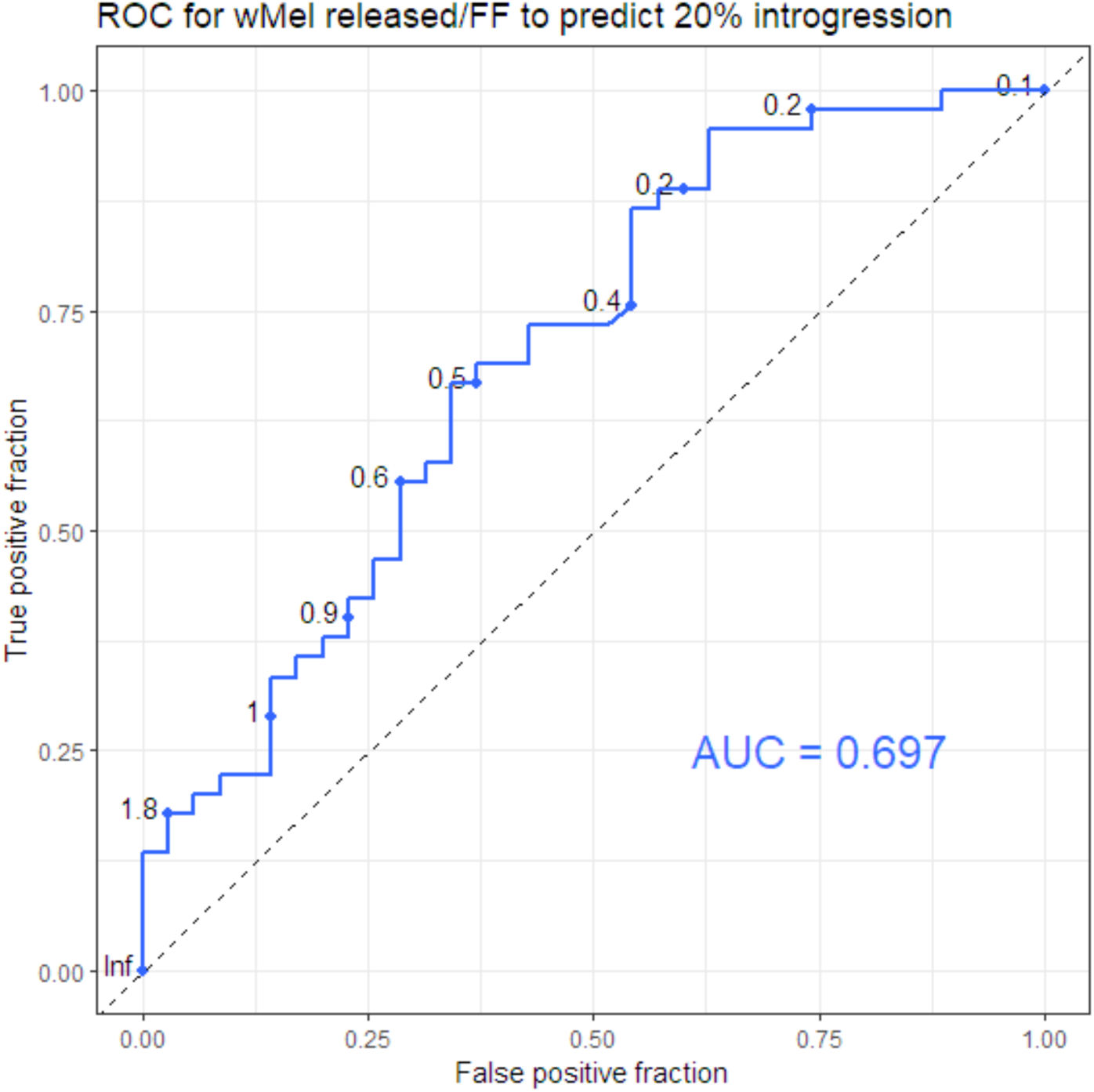
Receiver operating characteristic (ROC) curve showing classifying performance of the *w*Mel released/Fisher Ford Index to predict mean introgression at a 20% threshold.

**Figure S16.**
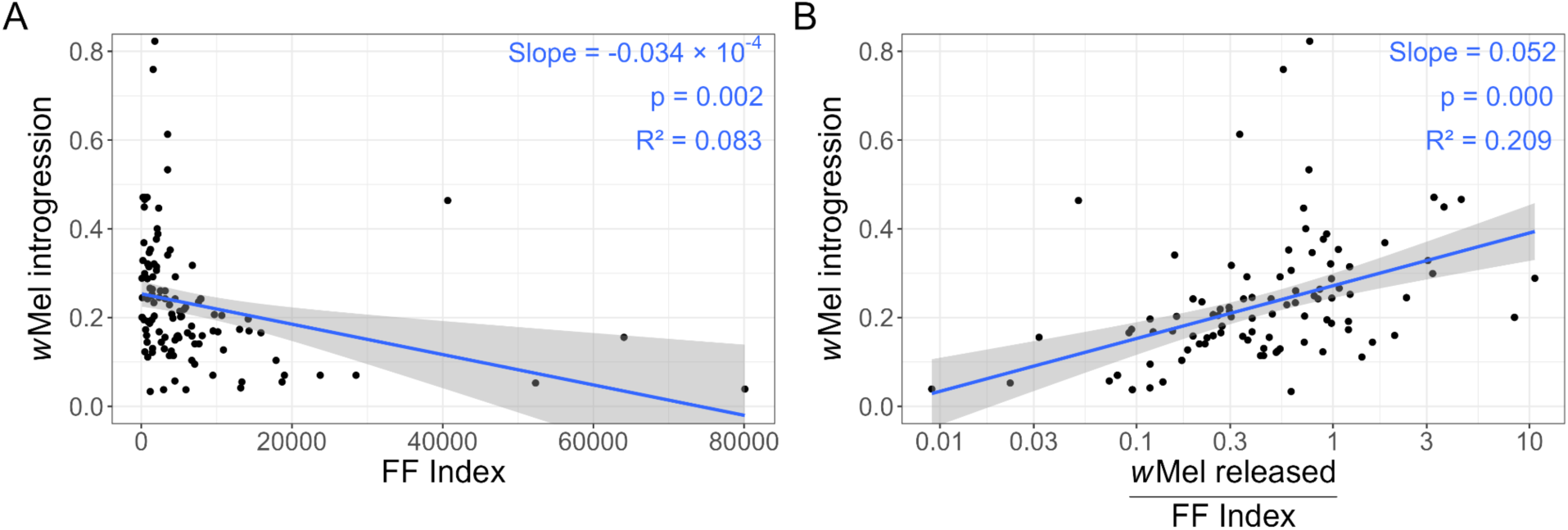
Baseline abundance estimates and introgression in traps with 9-36 days between first release and first trapping event, aggregated by 100m cells. Panels A and B show Fisher-Ford (FF) Index and ratio of wMel released (mean of releases before first trapping event) to Fisher-Ford Index versus mean *w*Mel introgression (log-transformed) for 117 cells. Blue lines show the linear regression trend with 95% CI. Blue text indicates regression slope, significance, and explained deviance (R^2^).

## Supplementary Tables

**Table S1.** Characteristics of cells by quartile of cumulative releases per cell.

|  | Quartile 1<br>(1262-6854 *w*Mel/cell)<br>N = 1,661 | Quartile 2<br>(6855-10428 *w*Mel/cell)<br>N = 1,661 | Quartile 3<br>(10429-14740 *w*Mel/cell)<br>N = 1,682 | Quartile 4<br>(14741-70074 *w*Mel/cell)<br>N = 1,637 |
| --- | --- | --- | --- | --- |
| **Intervention** | **Median (IQR)** |  |  |  |
| *w*Mel introgression (%) | 31 (16, 50) | 27 (15, 44) | 24 (14, 42) | 21 (12, 32) |
| Trap count | 6.1 (3.6, 9.1) | 5.5 (3.2, 9.3) | 5.2 (3.2, 9.7) | 6.2 (3.7, 9.6) |
| **Socioeconomic** |  |  |  |  |
| Inhabitants (0.01 km<sup>2</sup>) | 85 (38, 136) | 119 (71, 205) | 110 (65, 171) | 101 (60, 137) |
| Income (R\$/m) | 1,229 (829, 2,364) | 1,117 (768, 1,655) | 1,141 (770, 1,655) | 1,533 (1,205, 1,782) |
| **Environmental** |  |  |  |  |
| Impervious surface (%) | 0.59 (0.35, 0.79) | 0.74 (0.57, 0.85) | 0.76 (0.62, 0.86) | 0.80 (0.70, 0.87) |
| Green space (%) | 0.018 (0.003, 0.056) | 0.005 (0.000, 0.021) | 0.003 (0.000, 0.014) | 0.002 (0.000, 0.008) |
| Elevation (m) | 13 (8, 30) | 16 (9, 37) | 12 (8, 24) | 14 (9, 21) |

**Table S2.** Characteristics of cells by quartile of cumulative releases per inhabitant.

|  | Quartile 1<br>(0-21 *w*Mel per inhabitant)<br>N = 1,898 | Quartile 2<br>(22-79 *w*Mel per<br>inhabitant)<br>N = 1,898 | Quartile 3<br>(80-171 *w*Mel per<br>inhabitant)<br>N = 1,897 | Quartile 4<br>(172-5714 *w*Mel per<br>inhabitant)<br>N = 1,898 |
| --- | --- | --- | --- | --- |
| **Intervention** | **Median (IQR)** |  |  |  |
| *w*Mel introgression (%) | 34 (12, 52) | 22 (14, 38) | 26 (16, 42) | 24 (13, 40) |
| Trap count | 5.4 (3.1, 8.4) | 5.4 (3.2, 8.6) | 5.8 (3.4, 9.5) | 5.7 (3.5, 10.3) |
| **Socioeconomic** |  |  |  |  |
| Inhabitants (0.01 km<sup>2</sup>) | 69 (17, 163) | 160 (115, 236) | 105 (80, 131) | 41 (20, 72) |
| Income (R\$/m) | 1,297 (770, 2,631) | 1,021 (741, 1,555) | 1,492 (1,096, 1,928) | 1,385 (1,031, 1,754) |
| **Environmental** |  |  |  |  |
| Impervious surface (%) | 0.55 (0.18, 0.78) | 0.75 (0.60, 0.85) | 0.74 (0.61, 0.83) | 0.76 (0.54, 0.86) |
| Green space (%) | 0.02 (0.00, 0.08) | 0.00 (0.00, 0.02) | 0.00 (0.00, 0.02) | 0.00 (0.00, 0.02) |
| Elevation (m) | 14 (8, 32) | 22 (9, 43) | 16 (10, 25) | 11 (8, 17) |

**Table S3.**
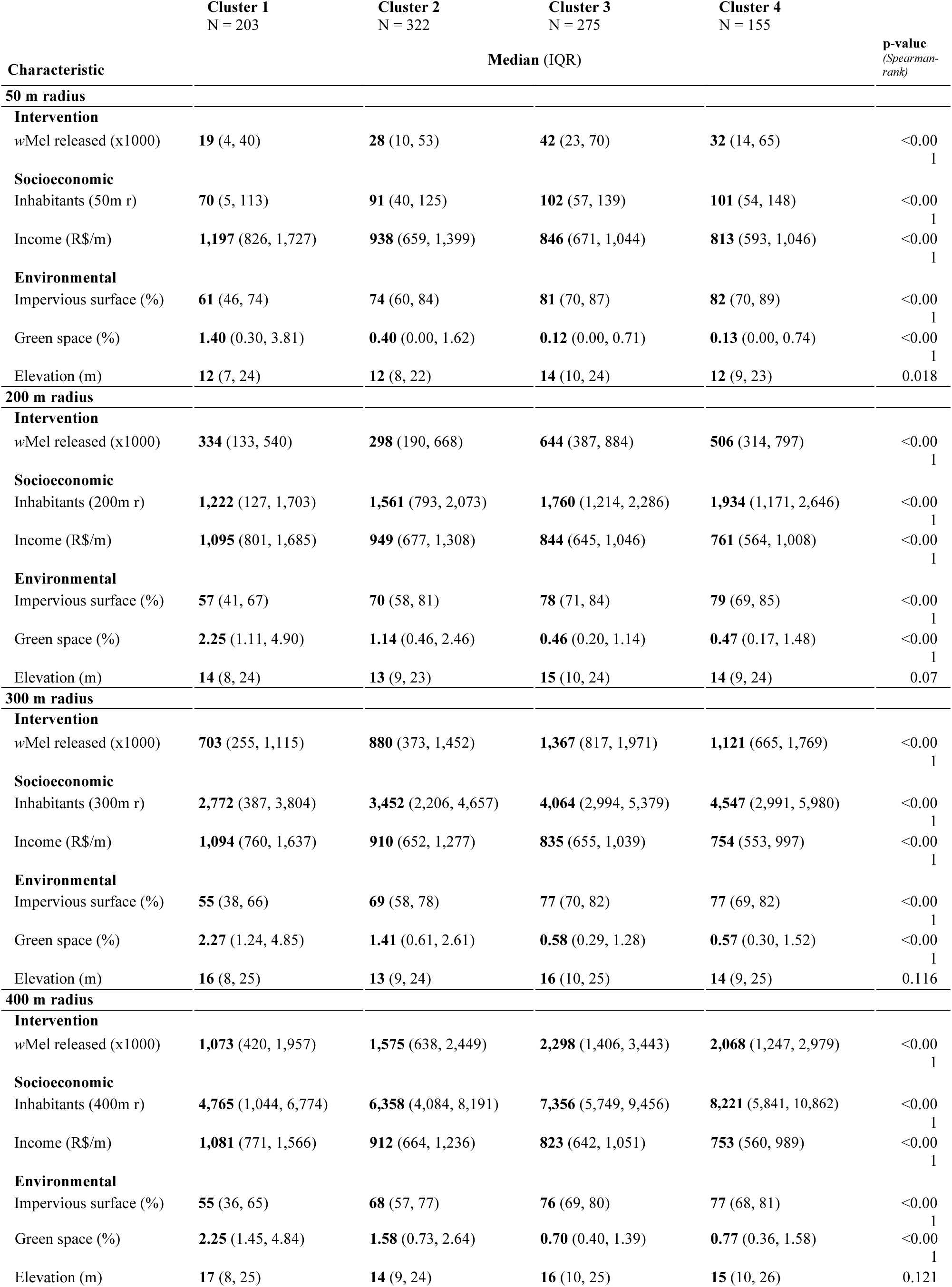
Characterization of GBMT clusters with covariates summarized by different radii around traps.

**Table S4.**
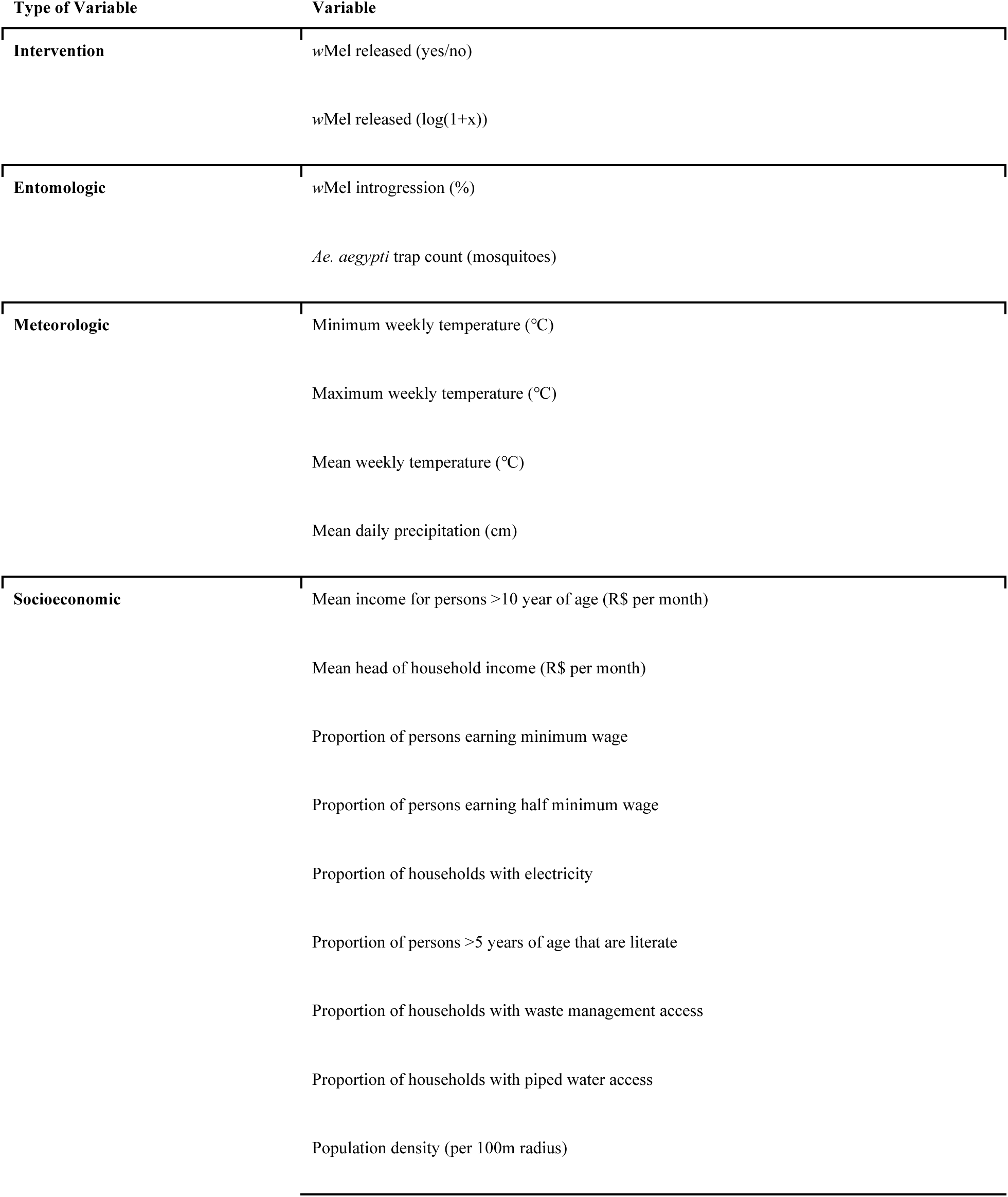
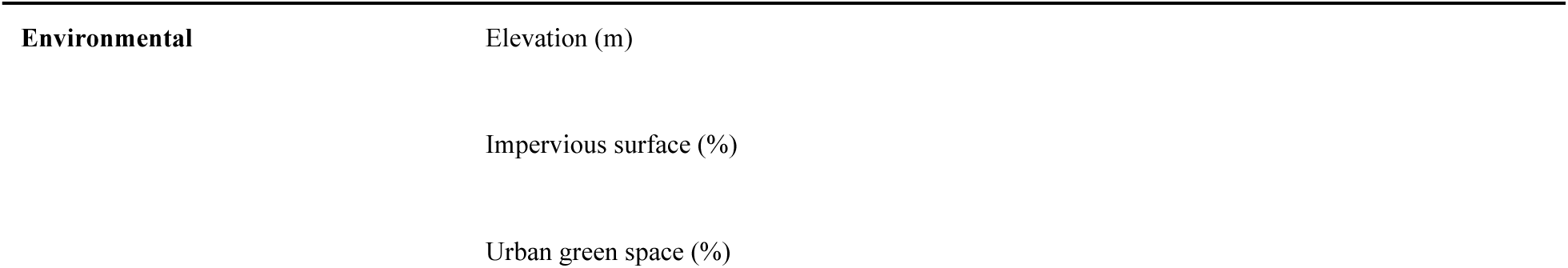
Variables tested as covariates in spatiotemporal models.

